# *TP53-*mediated clonal hematopoiesis confers increased risk for incident peripheral artery disease

**DOI:** 10.1101/2021.08.22.21262430

**Authors:** Seyedeh M. Zekavat, Vanesa Viana-Huete, Saman Doroodgar Jorshery, María A. Zuriaga, Md Mesbah Uddin, Mark Trinder, Kaavya Paruchuri, Nuria Matesanz, Virginia Zorita, Alba Ferrer-Pérez, Marta Amorós-Pérez, Scott M. Damrauer, Christie M. Ballantyne, Abhishek Niroula, Christopher J. Gibson, James Pirruccello, Gabriel Griffin, Benjamin L. Ebert, Peter Libby, Valentín Fuster, Hongyu Zhao, Marzyeh Ghassemi, Pradeep Natarajan, Alexander G. Bick, José J Fuster, Derek Klarin

## Abstract

**Background:** Somatic mutations in blood indicative of clonal hematopoiesis of indeterminate potential (CHIP), particularly in *DNMT3A*, *TET2*, and *JAK2*, are associated with an increased risk of hematologic malignancy, coronary artery disease, and all-cause mortality. However, whether CHIP is associated with increased risk of peripheral artery disease (PAD) remains unknown. In addition, chemotherapy frequently causes mutations in DNA Damage Repair (DDR) genes *TP53* and *PPM1D*, and whether CHIP caused by somatic mutations in DDR genes results in increased risk of atherosclerosis is unclear. We sought to test whether CHIP, and CHIP caused by DDR genes, associates with incident peripheral artery disease (PAD) and atherosclerosis.

**Methods:** We identified CHIP among 50,122 exome sequences in individuals from UK and Mass General Brigham Biobanks and tested CHIP status (N=2,851) with incident PAD and atherosclerosis across multiple arterial beds. To mimic the human scenario of clonal hematopoiesis and test whether the expansion of p53-deficient hematopoietic cells contributes to atherosclerosis, a competitive bone marrow transplantation (BMT) strategy was used to generate atherosclerosis-prone *Ldlr*-/- chimeric mice carrying 20% *Trp53*-/- hematopoietic cells (20% KO-BMT mice). We then evaluated aortic plaque burden and plaque macrophage accumulation 12 weeks after grafting.

**Results:** CHIP associated with incident PAD (HR 1.7; P=2.2×10^-5^) and atherosclerosis in multiple beds (HR 1.3; P=9.7×10^-5^), with increased risk among individuals with DDR CHIP (HR 2.0; P=0.0084). Among atherosclerosis-prone *Ldlr* null mice, the p53 -/- 20% KO-BMT mice demonstrated increased aortic plaque size (p=0.013) and accumulation of p53-/- plaque macrophages (P<0.001), driven by an abundance of p53-deficient plaque macrophages. The expansion of p53-deficient cells did not affect the expression of the pro-inflammatory cytokines IL-6 and IL-1β in the atherosclerotic aortic wall.

**Conclusions:** Our findings highlight the role of CHIP as a broad driver of atherosclerosis across the entire arterial system, with evidence of increased plaque among p53 -/- 20% KO-BMT mice via expansion of plaque macrophages. These observations provide new insight into the link between CHIP and cardiovascular disease, and lend human genetic support to the concept that post-cytotoxic chemotherapy patients may benefit from surveillance for atherosclerotic conditions in addition to therapy-related myeloid neoplasms.

## Introduction

Peripheral artery disease (PAD) is a leading cause of cardiovascular morbidity and mortality worldwide, and age is among its strongest risk factors. PAD associates with an extremely high cardiovascular mortality and unmitigated can progress to limb loss^1^. The age-related acquisition and expansion of leukemogenic mutations in hematopoietic stem cells has recently been associated with an increased risk of hematologic malignancy, coronary artery disease, and overall mortality^2, 3^. This phenomenon, termed clonal hematopoiesis of indeterminate potential (CHIP), is relatively common in asymptomatic older adults, affecting at least 10% of individuals older than 70 years of age^4^. CHIP mutations most frequently occur in epigenetic regulators *DNMT3A* and *TET2,* in DNA damage repair (DDR) genes *PPM1D* and *TP53*, or cell cycle and transcriptional regulator genes *JAK2* and *ASXL1*^5^. CHIP associates with coronary artery disease in multiple studies^2, 6^. Whether CHIP links with increased risk of atherosclerosis in the peripheral arterial bed (PAD) is unknown.

Here, we leveraged 50,122 whole exome sequences from two genetic biobanks (UK Biobank [UKB], Mass General Brigham Biobank [MGBB]) and tested whether CHIP was associated with increased risk of PAD and atherosclerosis across multiple arterial beds, and additionally whether these associations varied by putative CHIP driver gene. Based on these results, we then performed functional analyses in *Ldlr*-null mice transplanted with 20% *Trp53*-/- bone marrow cells, a murine model of atherosclerosis and clonal hematopoiesis driven by *TP53* mutations.

## Methods

### Cohorts and exclusion criteria

The UKB is a population-based cohort of approximately 500,000 participants recruited from 2006-2010 with existing genomic and longitudinal phenotypic data and median 10-year follow-up^7^. Baseline assessments were conducted at 22 assessment centres across the UK with sample collections including blood-derived DNA. Of ∼49,960 individuals with WES data available, we analyzed 37,657 participants consenting to genetic analyses after our exclusion criteria. Use of the data was approved by the Massachusetts General Hospital Institutional Review Board (protocol 2013P001840) and facilitated through UK Biobank Application 7089.

The MGBB contains genotypic and clinical data from >105,000 patients who consented to broad-based research across 7 regional hospitals and median 3-year follow-up^8^. Baseline phenotypes were ascertained from the electronic medical record and surveys. We analyzed 12,465 individuals consenting to genetic analysis after our exclusion criteria. Use of the data was approved by the Massachusetts General Hospital Institutional Review Board (protocol 2020P000904).

Across both cohorts, we excluded individuals with prevalent hematologic cancer, individuals without genotypic-phenotypic sex concordance, and one of each pair of 1^st^ or 2^nd^ degree relatives at random. For the UKB, samples were further restricted to individuals with Townsend deprivation index, a marker of socioeconomic status, available for analysis. Follow-up time was defined as time from enrollment to disease diagnosis for cases, or to censorship or death for controls.

### Whole exome sequencing and CHIP calling

UKB WES were generated from whole blood-derived DNA at the Regeneron Sequencing Center^9^. MGBB WES were generated using whole blood-derived DNA using Illumina sequencing (mean coverage 55x). Somatic CHIP variants were detected with GATK MuTect2 software with parameters as previously described^6, 10^. Common germline variants and sequencing artifacts were excluded as before. Samples were annotated with the presence of any CHIP if MuTect2 identified one or more of a pre-specified list of pathogenic somatic variants, as previously described^11, 12^. Additionally, samples were annotated with the presence of Large CHIP (variant allele frequency >10%), as larger CHIP clones have previously been more strongly associated with adverse clinical outcomes^6^.

### Phenotype definitions

Across both UKB and MGBB, PAD was defined by grouping together ICD-10 and ICD-9 billing codes for aortic atherosclerosis (I70), peripheral vascular disease (I73.8, I73.9), and operative procedures including amputation of leg (X09.3-5), bypass of artery of leg (L21.6, L51.3, L51.6, L51.8, L59.1-8), endarterectomy or angioplasty of leg artery (L52.1-2, L54.1,4,8, L60.1-2, L63.1,5), and other transluminal operations on leg arteries or peripheral stent placement (L63.9,L66.7), as previously described^13^. Additionally, in the UKB self-reported peripheral vascular disease, leg claudication/intermittent claudication, arterial embolism, femoral-popliteal leg artery bypass, leg artery angioplasty +/-stent, or amputation of leg were also incorporated (Data fields 20002, 20004) as performed previously^13^. Coronary artery disease and cerebral atherosclerosis phenotype definitions for each cohort are detailed in **Tables I** and **II in the supplement**. Other atherosclerotic conditions were defined using the Phecode Map 1.2^14^ ICD-9 (https://phewascatalog.org/phecodes) and ICD-10 (https://phewascatalog.org/phecodes_icd10) phenotype groupings for “abdominal aortic aneurysm”, “aortic aneurysm”, “other aneurysm”, “chronic vascular insufficiency of intestine”, “acute vascular insufficiency of intestine”, “atherosclerosis of renal artery”. The composite atherosclerosis phenotype was created by combining all analyzed atherosclerosis phenotypes (including PAD, coronary artery disease, cerebral atherosclerosis, abdominal aortic aneurysm, aortic aneurysm, other aneurysm, chronic vascular insufficiency of intestine, acute vascular insufficiency of intestine, and atherosclerosis of renal artery) into one phenotype, whereby the first instance across all of these phenotypes was used to determine time of first diagnosed atherosclerotic disease for survival analysis. Other phenotypic covariates (never/prior/current smoking status, hypertension, hyperlipidemia, principal components of ancestry, etc.) were used as previously defined^15^.

### Association Analysis

In the UKB and MGBB a traditional cox-proportional hazards model was utilized using the Survival package in R-3.5 adjusting for age, age^2^, sex, smoking status, normalized Townsend deprivation index as a marker of socioeconomic status (only available in UKB), and the first ten principal components of genetic ancestry. Demographic and clinical characteristics found to differ between individuals with and without CHIP were tested using chi-squared (categorical) and Wilcoxon-rank sum (continuous) tests with a two-tailed P< 0.05 determining significance. Sensitivity analysis including other covariates (normalized BMI, prevalent hypertension, prevalent Type 2 diabetes, and prevalent hyperlipidemia) was not found to significantly change associations with PAD in the UKB (**Figure I in the supplement**). Additional sensitivity analyses were utilized in the UKB including propensity score adjustment, as well as a marginal structural cox proportional hazards model estimated through stabilized inverse-probability-treatment-weight (IPTW)^16^ to estimate the total causal effect of CHIP on PAD. Further details of propensity score methods and stabilized IPTW analysis are described in the supplementary appendix. For our primary outcome PAD, results were combined across the UKB and MGBB using an inverse-variance weighted fixed effects meta-analysis, a two tailed association P < 0.05 determined statistical significance. In our secondary analysis of CHIP with 10 additional, incident atherosclerotic diseases, a two-tailed Bonferroni p-value threshold of P<0.05/10=0.005 was used to declare statistical significance.

### Mice

Animal experiments followed protocols approved by the Institutional Ethics Committee at the Centro Nacional de Investigaciones Cardiovasculares and conformed to EU Directive 86/609/EEC and Recommendation 2007/526/EC regarding the protection of animals used for experimental and other scientific purposes, enforced in Spanish law under Real Decreto 1201/2005. All mice were maintained on a 12-h light/dark schedule in a specific pathogen-free animal facility in individually ventilated cages and given food and water ad libitum. C57Bl/6J *Trp53*-/- mice were obtained from the Jackson Laboratory. *Ldlr*-/- mice carrying the CD45.1 isoform of the CD45 hematopoietic antigen were generated by crossing LDLR-KO mice from the Jackson Laboratory and B6.SJL-*PtprcaPepcb*/BoyCrl mice from Charles River Laboratories.

### Competitive bone marrow transplantation and atherosclerosis induction in mice

CD45.1+ *Ldlr*-/- recipients were transplanted with suspensions of BM cells containing 20% CD45.2+ *Trp53*-/- cells and 80% CD45.1+ *Trp53*+/+ cells (20% KO-BMT mice) or 20 % CD45.2+ *Trp53*+/+ cells and 80% CD45.1+ *Trp53*+/+ cells (20% WT-BMT mice) similar to previous studies^17^ (**Figure II in the supplement**). BM cells were isolated from femurs and tibias of donor mice after euthanasia. Recipient *Ldlr*-/- mice were exposed to two doses of 450 rad three hours apart. After the second irradiation, each recipient mouse was injected with 10^7^ BM cells i.v. Water was supplemented with antibiotics for 7 days before transplant and for 14 days post-transplant. Mice that did not recover full pre-irradiation body weight 28 days after transplant were excluded from further analysis. Starting four weeks after BMT, mice were fed a high fat high cholesterol (HF/HC) Western diet (Harlan-Teklad, TD.88137, Adjusted Calories Diet; 42% from fat, 0.2% cholesterol) to promote hypercholesterolemia and the development of atherosclerosis. Mice were maintained on a HF/HC diet for 9 weeks

### Quantification of aortic atherosclerosis burden in mice

Mice were euthanized and aortas were removed after in situ perfusion with phosphate-buffered saline injected through the left ventricle of the heart. Tissue fixation was achieved by immersion in 4% paraformaldehyde in PBS overnight at 4°C. Aortic tissue was then dehydrated and embedded in paraffin for sectioning. Histological sections comprising the aortic root as determined by the location of the aortic valve leaflets were cut at a thickness of 4 µm or 6 µm. An operator who was blinded to genotype quantified plaque size in aortic root sections by computer-assisted morphometric analysis of microscopy images. For each mouse, atherosclerotic plaque size in aortic root cross-sections was calculated as the average of 5 independent sections separated by ∼16µm. Atherosclerotic plaque composition was examined by immunohistochemical techniques. Vascular smooth muscle cells were identified with an alkaline phosphatase-conjugated mouse anti-smooth muscle α-actin (SMA) monoclonal antibody (clone 1A4, SIGMA) and Vector Red Alkaline Phosphatase Substrate (Vector Laboratories). Macrophages were detected with a rabbit anti-Mac2 monoclonal antibody (Santa Cruz Biotechnologies), a biotin-conjugated goat anti-rat secondary antibody and streptavidin-HRP, DAB substrate (all from Vector Laboratories), with hematoxylin counterstaining. Collagen content was determined by a modified Masson’s trichrome staining. Microscopy images were analyzed using ImageJ software using the Color Deconvolution plugin. Plasma was collected in EDTA-coated tubes and cholesterol levels were determined using an enzymatic assay (Cholesterol E, WAKO Diagnostics).

### Flow cytometry analyses of blood and tissue samples

Peripheral blood was obtained from the facial vein and collected into EDTA-coated tubes. Bone marrow cells were flushed out of two femurs and two tibias per mouse. Aortic arches were digested for 45 minutes at 37°C in RPMI containing 10% FBS and 0.25 mg/ml Liberase TM (Roche Life Science). Red blood cells were lysed in all samples by treatment with 1X Red Blood Cell Lysis Buffer (eBioscience ThermoFisher) for 5 minutes on ice. Bone Marrow lineage-negative cells were defined as negative for CD11b, Gr-1, Ter119, B220, CD3e and CD127. Bone marrow HSPCs were defined as Lineage-, c-Kit+, Sca1+. Blood classical monocytes were identified as CD45+, CD115^Hi^, CD43^Lo^, Ly6c^Hi^; patrolling monocytes, as CD45+, CD115^Hi^, CD43^Hi^, Ly6c^Lo^; neutrophils, as CD45+, CD115^Int^, Ly6g+; T lymphocytes, as CD45+, CD115-, B220-, CD3+; B lymphocytes, as CD45+, CD115-, CD3-, B220+; and aortic macrophages, as CD45+, CD3-, Ly6g-, CD11B+, F4/80Hi. Gating strategies can be found in our previous publications^17, 18^. Samples were stained with combinations of biotinylated and/or fluorescently labeled antibodies in PBS with 1% FBS for 30 min on ice. The following fluorescent antibodies were used for staining and flow cytometry analysis: eFluor450-conjugated anti-CD45.2, FITC-conjugated anti-CD45.2, Pe-Cy7-conjugated anti-CD45.1, FITC-conjugated anti-CD11b, eFluor450-conjugated anti-CD11B, PE-conjugated anti-CD115, PE-eFluor610-conjugated anti-CD3, FITC-conjugated anti-CD4 (from eBioscience); APC-Cy7-conjugated anti-B220, PerCP-Cy5.5-conjugated anti-Ly6G, BV510-conjugated anti-Ly6G, BV711-conjugated anti-CD43, PE-Cy7-conjugated anti-c-Kit, AlexaFluor647-conjugated anti-Sca-1 (from BD Biosciences); PE-conjugated anti-F4/80 (from R&D Systems); PerCP-Cy5.5-conjugated anti-CD45.1, BV510-conjugated anti-CD8a, PerCP-Cy5.5-conjugated anti-mouse Ki-67 (from BioLegend). Fixation/permeabilization for Ki-67 intracellular staining was achieved using commercially available kits following manufacturer’s instructions (Foxp3 Transcription Factor Staining Buffer Set, eBioscience ThermoFisher). Dead cells were excluded from analysis by DAPI staining in unfixed samples and by LIVE/DEAD Fixable Near-IR staining (ThermoFisher Scientific) in fixed samples. BD LSRFortessa and BD FACSymphony Cytometers (BD Bioscience) were used for data acquisition. Data were analyzed with FlowJo Software.

### Murine macrophage culture and cell cycle analysis

Bone marrow-derived macrophages (BMDM) were obtained from suspensions of femoral BM and differentiated for 7 days in the presence of RPMI Medium supplemented with antibiotics, 10% fetal bovine serum and 100 ng/ml MCSF. For cell-cycle analysis, MCSF concentration in cell culture medium was decreased to 5 ng/ml for 48h to induce progressive synchronization of macrophages in G0 phase (i.e. quiescence) and cell cycle re-entry was induced by treatment with 100 ng/ml MCSF. Macrophages were trypsinized and collected by centrifugation for 5 minutes at 300g. After fixation in 80% ethanol for 1 h at −20°C, cells were incubated for at least 30 minutes with 50 µg/mL propidium iodide containing 0.25 mg/mL RNAse A (both from SIGMA). Labelled cells were analyzed in a BD LSRFortessa flow cytometer BD Bioscience and DNA histograms were fitted into cell cycle distributions using ModFit 3.0 software (Verity Software House).

### Gene expression analysis by quantitative real-time PCR (qPCR) and Western Blot

Total RNA from aortic arch tissue was isolated using Trizol reagent and RNeasy kits (QIAGEN). RNA (1 µg) was reverse transcribed with High-Capacity cDNA Reverse Transcription Kit (Applied Biosystems) and qPCR was performed with SYBR® Green PCR Master Mix (Applied Biosystems) in a AB7900 Real time PCR system. Results were analyzed with the ΔΔCt method. The average of 36B4 and β-actin was used as reference for normalization. Primer sequences can be found in our previous publication^17^. For Western Blot analysis of protein levels, protein extracts from cultured macrophages were obtained using ice-cold lysis buffer (20mM Tris-HCl pH 7.5, 150 mM NaC, 1 mM NA2EDTA, 1 mM EGTA, 1% Triton) supplemented with protease and phosphatase inhibitors (Roche Applied Science). Equal amounts of protein lysates were resolved by SDS-PAGE and the following antibodies were used for immunoblotting: anti-p53 (Cell Signaling, #32532S, dil 1/1000) and HRP anti-alpha tubulin (AbCam, #ab40742, dil 1/2000). An ImageQuant LAS 4000 biomolecular imaging system (GE Healthcare) was used for image acquisition and the ImageJ software was used for band densitometric analysis.

### Statistical analysis of data in experimental studies

Data are shown as mean ± SEM unless otherwise stated. Statistical significance of differences in experiments with two groups and only one variable was assessed by unpaired Student’s t tests (with Welch correction for unequal variance when appropriate) or Mann-Whitney U Tests. Differences in experiments with more than one independent variable were evaluated by two-way analysis of variance (ANOVA) with post-hoc Sidak’s multiple comparison tests. All statistical tests were performed using GraphPad Prism software (GraphPad Software Inc.).

## Results

### Study Cohorts and Risk Of Hematologic Malignancy

After excluding individuals with a known history of hematologic malignancy at enrollment, we identified 37,657 unrelated individuals from the UKB and 12,465 individuals from MGBB with whole exome sequencing data available for downstream analysis. Using a previously validated somatic variant detection algorithm^10^, we identified 2,194 (5.8%) and 657 (5.4%) CHIP carriers in the UKB and MGBB, respectively (**Table III in the supplement**). Demographic and clinical characteristics of these individuals, stratified by CHIP status, are depicted in **Table IV in the supplement**. CHIP carriers tended to be older, male, previous smokers, and have a history of coronary artery disease, hypertension, and hyperlipidemia (two-tailed chi-squared and Wilcoxon-rank sum P< 0.05).

We first replicated known CHIP associations^10^ with white blood cell (Beta 0.09 SD; 95% CI 0.05-0.13; P=1.6×10^-5^), monocyte (Beta 0.05 SD; 95% CI 0.01-0.09; P=0.009), neutrophil (Beta 0.10 SD; 95% CI 0.06-0.14; P=2.1×10^-6^), and platelet counts (Beta 0.07 SD; 95% CI 0.03-0.11; P=0.0005) in UKB, with larger CHIP clone size as measured by variant allele fraction (VAF) having stronger effects on blood counts (**Figure III in the supplement**). Consistent with the existing literature^5, 10^, CHIP also associated with incident hematologic malignancy (HR 2.20; 95% CI 1.70-2.85; P=1.8×10^-9^) - specifically for acute myeloid leukemia (HR 8.08; 95% CI 4.36-14.97; P=3.2×10^-11^), myeloproliferative neoplasms (HR 5.89; 95% CI 3.69-9.89; P=9.7×10^-14^), and polycythemia vera (HR 12.37; 95% CI 4.85-31.54; P=1.4×10^-7^). This risk increased with larger VAF (**Figure IV in the Supplement**).

### CHIP and Incident PAD Risk

We next tested the association of CHIP status with incident PAD. Using available electronic health record (EHR) data and a previously validated PAD definition^13^, we identified 338 and 419 incident PAD cases in UKB and MGBB, respectively. CHIP associated with a 58% increased risk of incident PAD in the UKB (HRUKB = 1.58, 95% CI: 1.11-2.25; P=0.01, **Figure 1**), results that were replicated in MGBB (Overall HR = 1.66, 95% CI: 1.31-2.11; P=2.4×10^-5^). We then sought to evaluate whether those with larger CHIP clone sizes (i.e., higher VAF) had greater risk for PAD, as larger CHIP clones associate more strongly with adverse clinical outcomes^6^. We observed a graded relationship between CHIP VAF and PAD, as those with a VAF > 10% had even greater risk for an incident PAD event (Overall HR = 1.97, 95% CI: 1.44-2.71; P=2.3×10^-5^, **Figure 1**). Additional sensitivity analyses, including propensity score adjustment and a marginal structural Cox proportional hazards model estimated through stabilized inverse-probability-treatment-weight revealed similar results in the UKB (**Figure V in the supplement**). Subsequent analyses showed no significant interaction between CHIP status and either age, sex, or smoking status on incident PAD risk.

**Figure 1.**
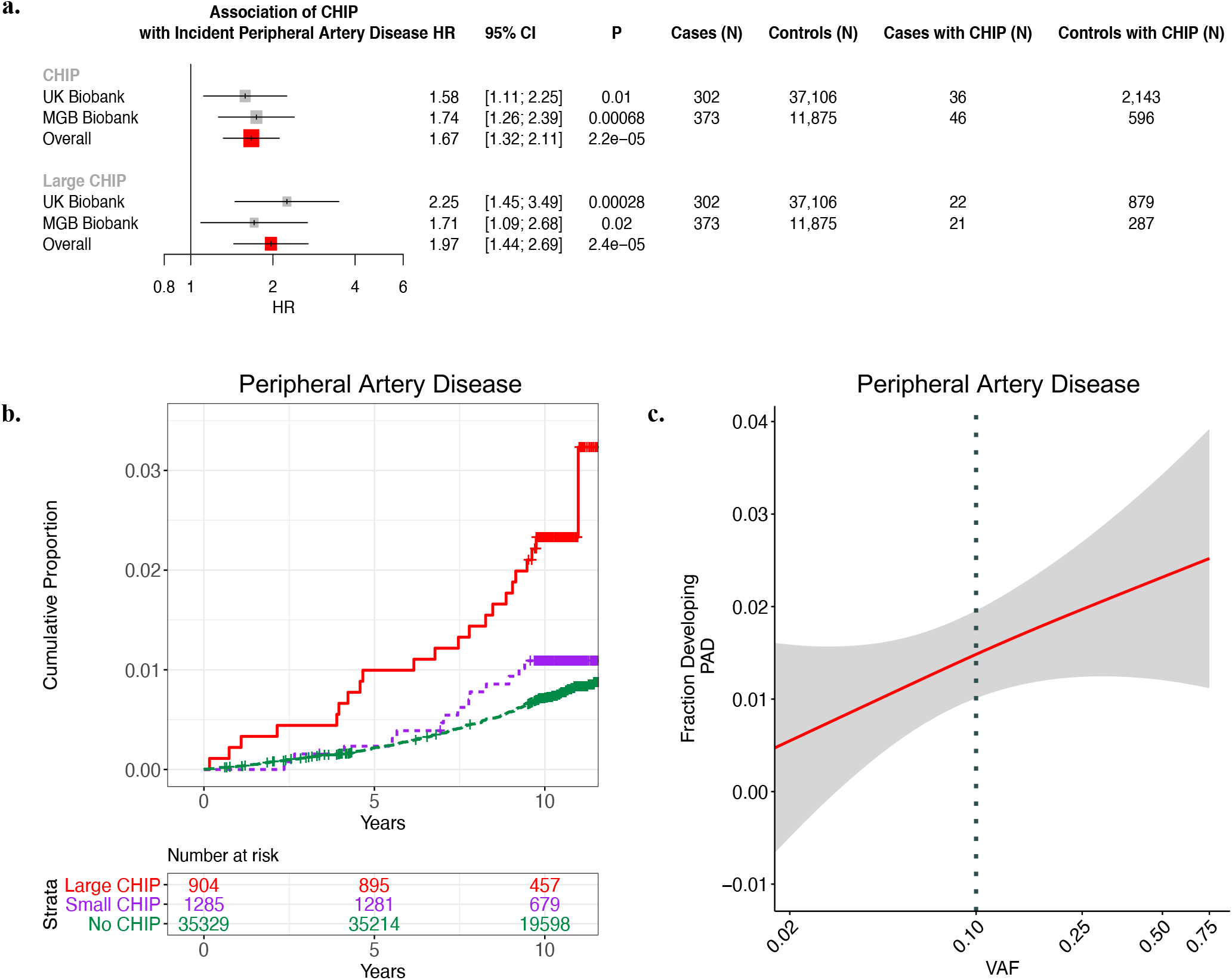
CHIP and incident PAD risk. a) Association of CHIP and large CHIP (VAF>10%) carrier state with incident PAD events in the UK Biobank (UKB) and Mass General Brigham Biobank (MGBB). Results were combined using an inverse-variance weighted fixed effects meta-analysis. b) Cumulative proportion of individuals developing PAD stratified by CHIP VAF clone size category in the UK Biobank. c) Fraction of individuals developing incident PAD by CHIP VAF in the UK Biobank. CHIP = clonal hematopoiesis of indeterminate potential; VAF = variant allele fraction; PAD = peripheral artery disease

### CHIP and Incident Atherosclerosis Across Multiple Vascular Beds

We next assessed whether CHIP was associated with 9 other incident atherosclerotic diseases across multiple vascular beds. Using EHR-based disease definitions^19^, we tested the association of CHIP with atherosclerotic disease across the mesenteric (acute and chronic), coronary, and cerebral vascular beds, as well as with aneurysmal disease (aortic and any other aneurysm). We observed significant associations for coronary artery disease (HR 1.40, 95% CI: 1.20 to 1.63; P=1.9×10^-5^), any aortic aneurysm (HR 1.74; 95% CI: 1.21 to 2.51; P=0.0028), other aneurysms (HR 1.70; 95% CI: 1.23 to 2.34; P=0.0013), and chronic mesenteric ischemia (HR; 95% CI: 2.34 to 35.63; P=0.0015) across both cohorts, with directionally consistent effect estimates observed for all the tested phenotypes (**Figure 2a**). These associations were consistently stronger for large CHIP clones (**Figure VI in the supplement)**. We then created a composite, incident atherosclerosis outcome combining all nine atherosclerotic phenotypes (“pan-arterial atherosclerosis”, **Table V in the Supplement**). CHIP associated with this combined incident pan-arterial atherosclerosis endpoint (HR 1.31, 95% CI: 1.14 to 1.49, P=9.7×10^-5^), again with stronger effects conferred by large CHIP clones (HR 1.45; 95% CI: 1.20 to 1.75; P=0.00013) (**Figure 2b,c**).

**Figure 2.**
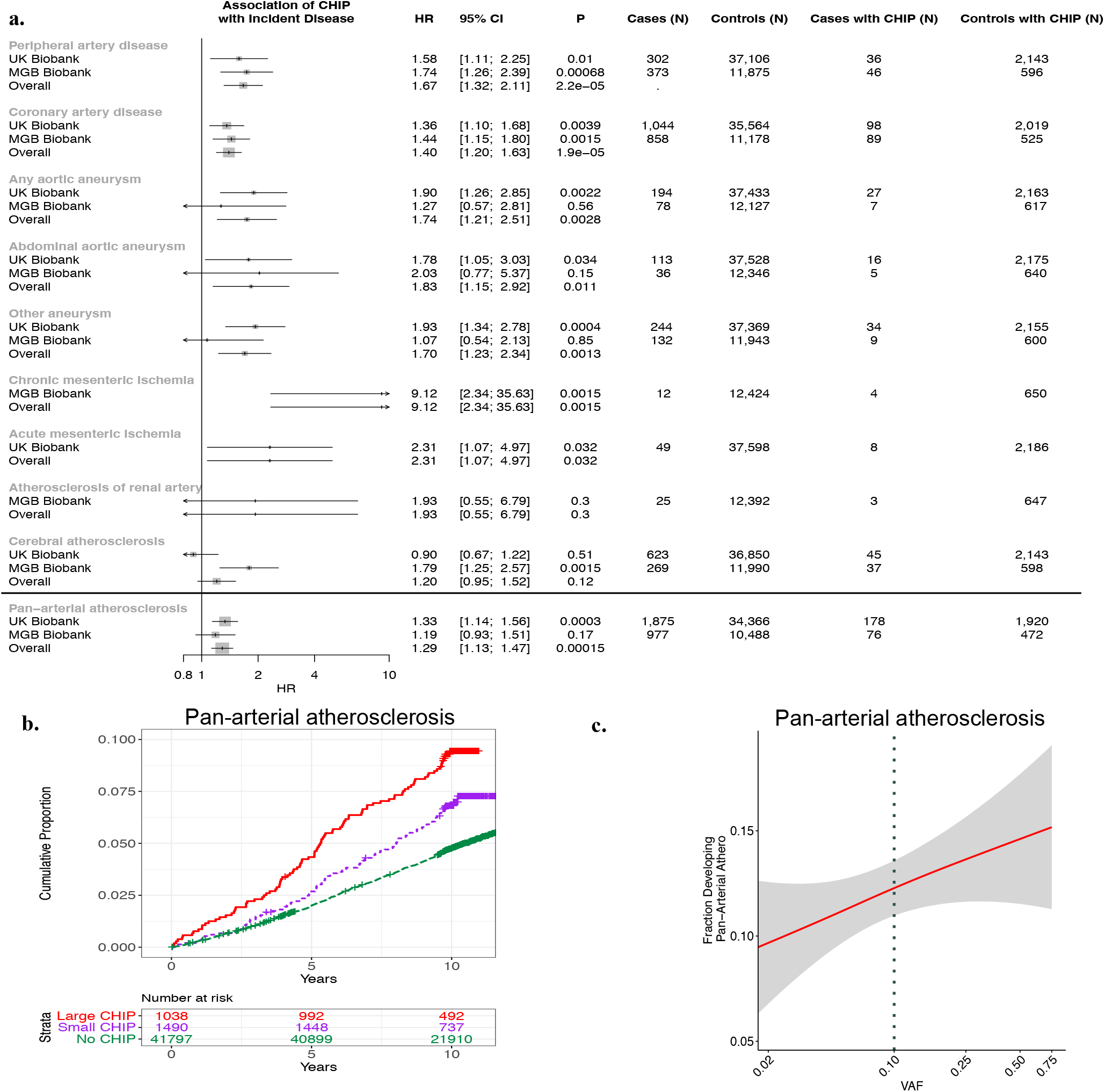
CHIP and incident pan-arterial atherosclerosis risk. a) Association of CHIP with 9 incident atherosclerotic diseases separately and combined in a ‘Pan-arterial atherosclerosis’ phenotype in the UKB, MGBB, and meta-analyzed across both studies (“Overall”). b) Cumulative risk of incident atherosclerosis across the composite ‘pan-arterial atherosclerosis’ phenotype stratified by no CHIP, small CHIP (VAF<10%), and large CHIP (VAF≥10%) carrier state in the UK Biobank. c) Association of CHIP VAF with fraction of individuals developing pan-arterial atherosclerosis in the UK Biobank. CHIP = clonal hematopoiesis of indeterminate potential; VAF = variant allele fraction; PAD = peripheral artery disease

### Gene-specific analyses of CHIP with incident atherosclerotic diseases

Next, we sought to understand whether the clonal hematopoiesis putative driver gene differentially affected the risk of acquiring atherosclerosis. Previous work has focused primarily on the epigenetic regulators *DNMT3A* and *TET2*^3, 17^, and whether DDR CHIP confers an increased risk of atherosclerosis is unknown. We stratified the CHIP-PAD and CHIP pan-arterial atherosclerosis analyses by putative driver genes and specific mutations - focusing on *DNMT3A*, *TET2*, *ASXL1*, *JAK2*, the DDR genes *PPM1D* and *TP53*, and mutations that specifically disrupt splicing factor genes *(LUC7L2, PRPF8, SF3B1, SRSF2, U2AF1,* and *ZRSR2)*^20^. We observed an association of CHIP with PAD across the four common CHIP genes (*DNMT3A*, *TET2*, *ASXL1*, and *JAK2*), with significant heterogeneity of incident PAD effect sizes across the CHIP genes (P_heterogeneity_ = 0.03) (**Figure 3a**). This heterogeneity persisted in sensitivity analysis after excluding *JAK2* carriers (P_heterogeneity_ = 0.046). These data also revealed the novel finding that DDR *TP53* and *PPM1D* CHIP associates with incident PAD (HR 2.72; 95% CI: 1.20 to 1.75; P=0.00013) and incident CAD (HR 2.51; 95% CI: 1.52-4.14; P=0.00032), with a stronger effect on PAD conferred by *TP53* (HR 4.98; 95% CI: 1.23-20.09; P=0.024, **Figure 3a-c).** Similar findings were observed for the incident pan-arterial atherosclerosis outcome when stratifying by putative driver gene (**Figure VII in the supplement**). Further sensitivity analysis for DDR-CHIP and incident PAD when excluding solid organ malignancy did not significantly change the associations (P_heterogeneity_ > 0.05).

**Figure 3:**
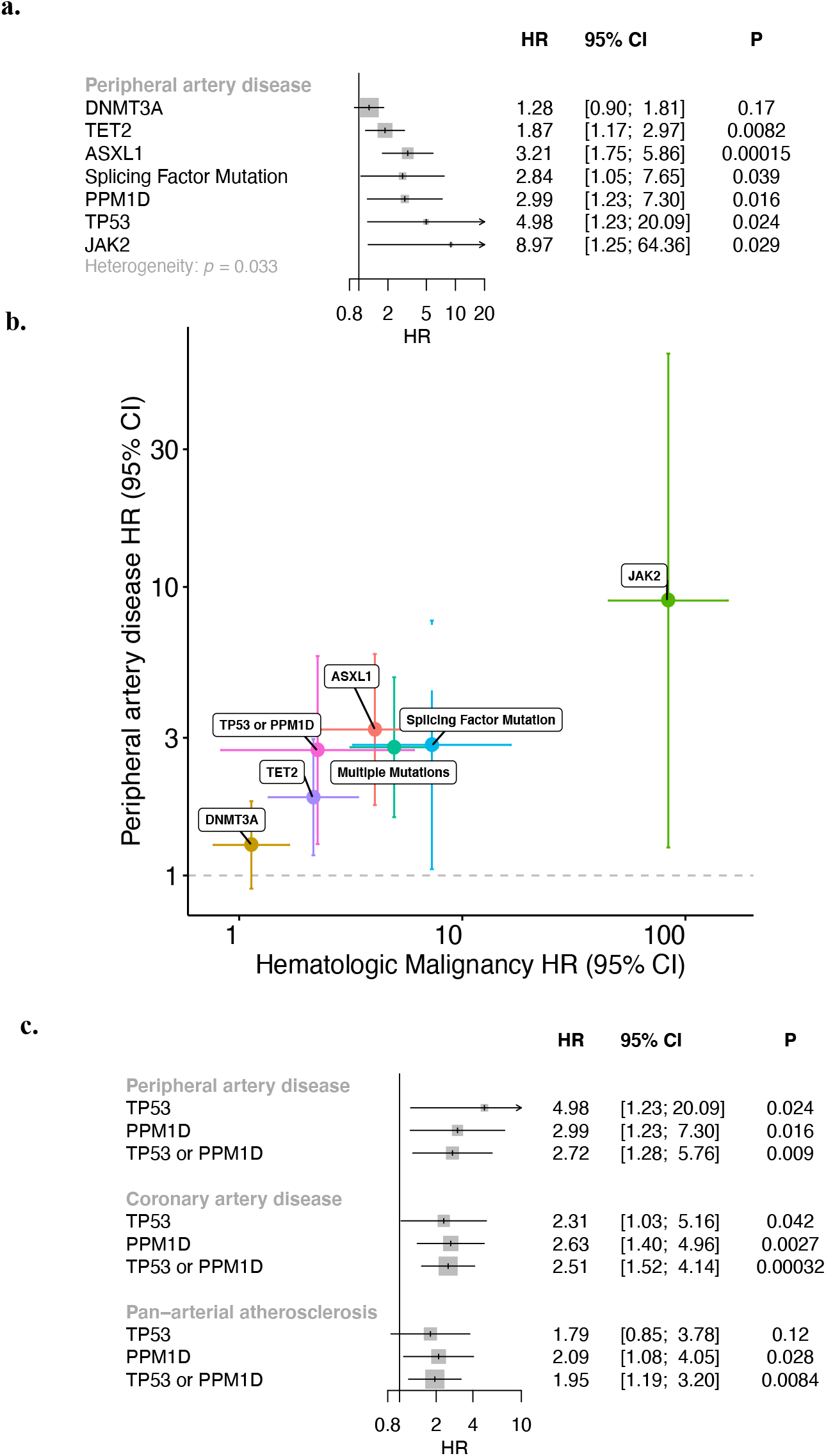
Gene-specific association of CHIP with incident peripheral artery disease (PAD). a) CHIP-PAD association analyses stratified by putative CHIP driver gene. Results following meta-analysis across the UKB and MGBB are shown. b) Gene-specific comparison of HR and 95% CI for hematologic malignancy (x-axis) and PAD (y-axis) in the UKB. c) Association of DDR CHIP (*PPM1D or TP53*) with incident peripheral artery disease, coronary artery disease, and pan-vascular atherosclerosis. Results across UK Biobank and MGB Biobank were combined using an inverse-variance weighted fixed effects meta-analysis. CHIP = clonal hematopoiesis of indeterminate potential; DDR = DNA-damage repair; VAF = variant allele fraction; PAD = peripheral artery disease

### Atherosclerosis development in p53-/ CHIP mice

Based on our gene specific findings, we next further characterized the effects of reduced function of hematopoietic p53 in atherosclerotic mice. To mimic CHIP and test whether the expansion of p53-deficient hematopoietic cells contributes to atherosclerosis, a competitive bone marrow transplantation (BMT) strategy was used to generate atherosclerosis-prone *Ldlr*-/- chimeric mice carrying 20% *Trp53*-/- hematopoietic cells (20% KO-BMT mice). These mice then consumed a high fat/high cholesterol diet for 9 weeks to induce atherosclerosis development. To distinguish donor *Trp53*-/- and *Trp53*+/+ cells in this experimental setting, *Trp53*+/+ cells were obtained from mice carrying the CD45.1 variant of the CD45 hematopoietic antigen, whereas *Trp53*-/- cells were obtained from mice carrying the CD45.2 variant of this protein. Control mice (20% WT-BMT) were transplanted with 20% CD45.2+ *Trp53*+/+ cells and 80% CD45.1+ *Trp53*+/+ cells (**Figure 4a and Figure II in the supplement**). Flow cytometry analysis of CD45.2+ blood cells established that this BMT strategy led to a modest, but significant expansion of donor *Trp53*-/- BM-derived cells compared to *Trp53*+/+ cells in both BM hematopoietic stem/progenitor cells (HSPCs) and circulating white blood cells (**Figure 4B,C**), consistent with previous studies^21–23^. Transplanted *Trp53*-/- BM cells expanded into all blood cell lineages to a similar extent, but this relative expansion did not affect absolute blood cell counts (**Figure VIII in the supplement**). Having validated this mouse model of p53 CHIP based on a competitive BMT strategy, we next assessed whether this phenomenon affects the development of atherosclerosis or related metabolic abnormalities. The presence and expansion of *Trp53*-/- cells led to a significant ∼40% increase in plaque size in the aortic root of male *Ldlr*-/- mice (**Figure 4D**), without affecting body weight, spleen weight or serum cholesterol levels (**Figure VIII in the supplement, C-E**). Similar results were obtained in female *Ldlr*-/- mice (**Figure IX in the supplement**). In contrast to previous work on *DNMT3A/TET2* CHIP, the expansion of p53- deficient cells^12, 17, 18, 24, 25^ did not affect the expression of the pro-inflammatory cytokines IL-6 and IL-1β in the atherosclerotic aortic wall (**Figure X in the supplement**).

**Figure 4:**
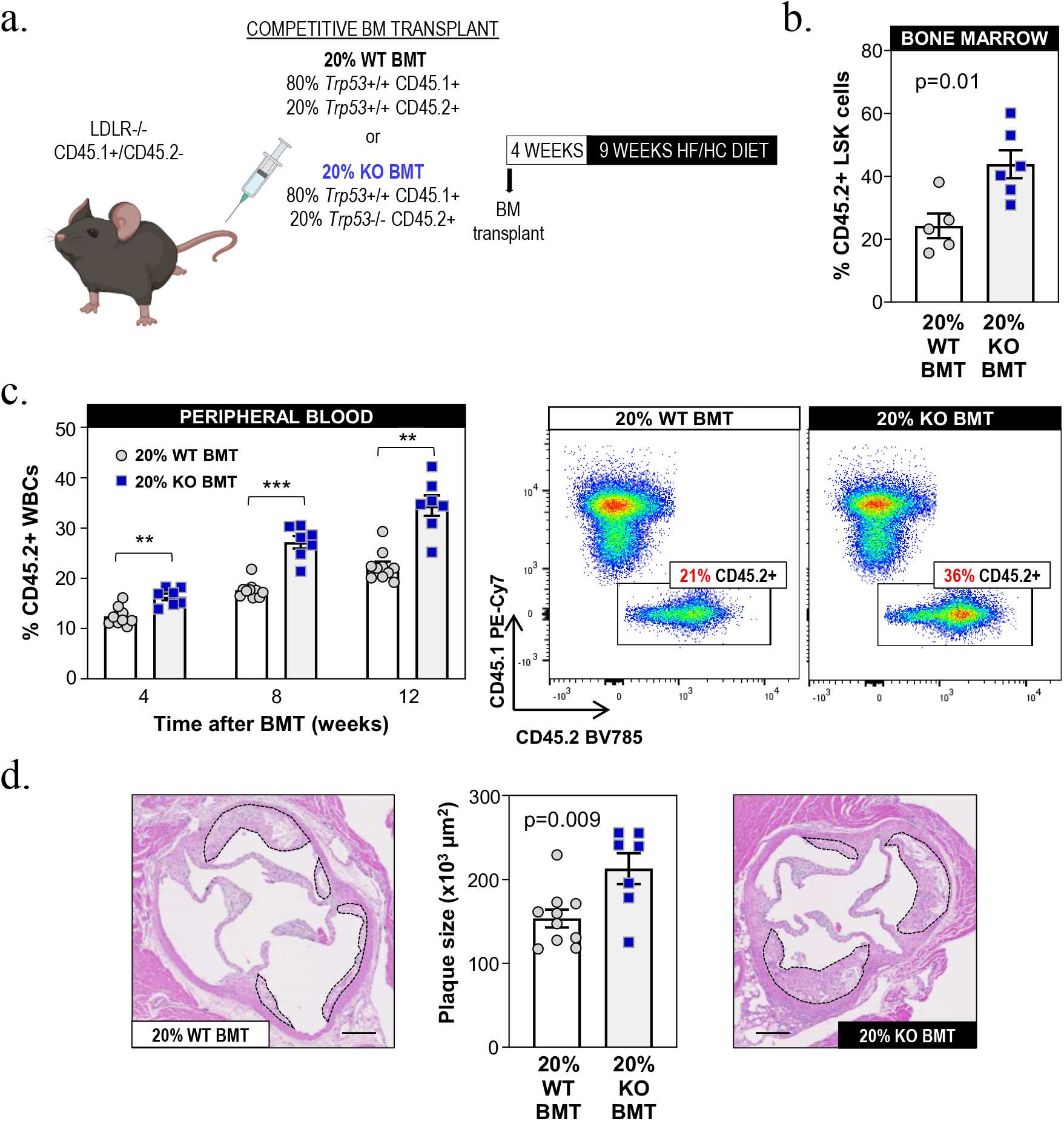
Accelerated atherosclerosis in a murine model of *TP53* mutation-driven CHIP. a) Summary of the competitive BMT approach and the timeline of these studies. 20% KO-BMT male mice and 20% WT-BMT controls were fed a high-fat/high-cholesterol (HF/HC) diet for 9 weeks, starting 4 weeks after BMT (n=10 20% WT-BMT, n=7 20% KO-BMT, unless otherwise noted). b) Percentage of CD45.2+ cells in the bone marrow HSPC population (Lin- Sca1+ cKit+ cells) after 9 weeks on HF/HC diet (13 weeks post-BMT), evaluated by flow cytometry (n= 5 20% WT-BMT, n=6 20% KO-BMT). c) Percentage of CD45.2+ in white blood cells, evaluated by flow cytometry (**p<0.01, ***p<0.001). Representative CD45.1/CD45.2 dot plots are shown. d) Aortic root plaque size. Representative images of hematoxylin and eosin-stained sections are shown; atherosclerotic plaques are delineated by dashed lines. Scale bars, 100 µm.

### Proliferation and expansion of p53-deficient macrophages in the murine atherosclerotic aorta

Increased atherogenesis in mice carrying *Trp53*-/- cells was paralleled by a substantial increase in plaque macrophage content, as assessed by immunohistological staining of Mac2, with no significant changes in other cell components (**Figure 5A**), suggesting a contribution of increased arterial macrophage burden to accelerated atherosclerosis in conditions of p53 CHIP. Flow cytometry analysis of matched samples from blood and digested atherosclerotic aortae from 20% KO-BMT mice revealed substantially higher chimerism in aortic macrophages (∼76% *Trp53*-/-) than in blood classical monocytes (∼48%), the major source of plaque macrophages (**Figure 5B**). This expansion of p53-deficient macrophages within the vascular wall was paralleled by a 2-fold increase in the frequency of proliferating cells within the CD45.2+ *Trp53*- /- aortic macrophage population compared to CD45.2+ *Trp53*+/+ macrophages, as assessed by flow cytometry analysis of the proliferation-related antigen Ki-67 (**Figure 5C)**. Consistent with our observations *in vivo*, cultured *Trp53*-/- macrophages exhibit accelerated mitotic cell cycle progression, with a >2-fold increase in the percentage of S-phase cells upon stimulation with macrophage colony stimulating factor (MCSF), a major determinant of plaque macrophage proliferation^26^ (**Figure 5D**). *Trp53* expression analysis suggests a central role of p53 in the normal regulation of macrophage cell cycle progression, as p53 was expressed in quiescent macrophages at the transcript and protein level and further induced after MCSF stimulation (**Figure 5E,F**). Furthermore, p53-deficient macrophages exhibited major changes in the expression of pivotal regulators of cell cycle entry and progression, such as *Cdkn1a*/p21^Cip1^ and Cyclin B1 (**Figure 5G**).

**Figure 5:**
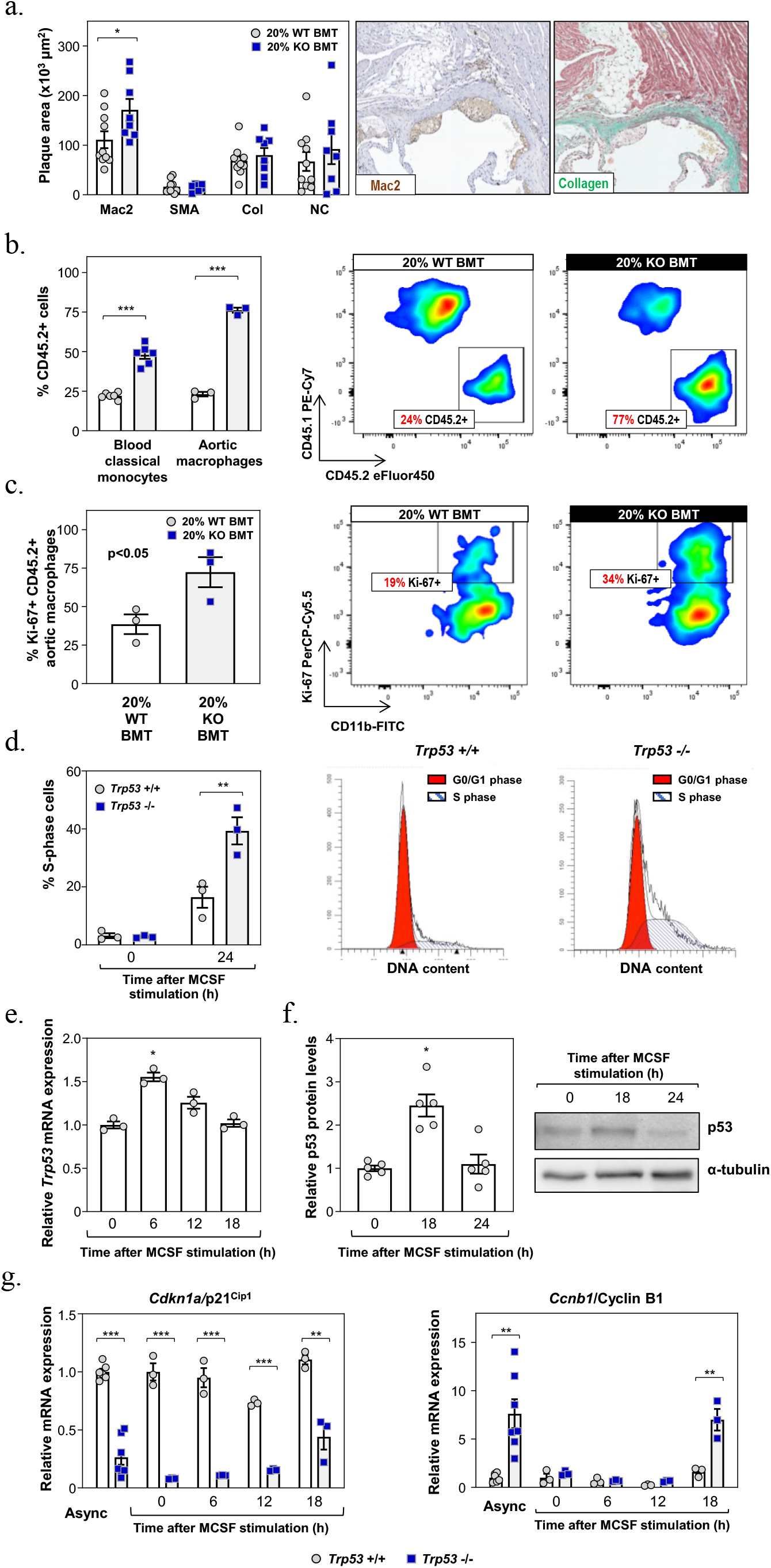
Increased proliferation and expansion of p53-deficient macrophages. a) Plaque composition in 20% KO BMT female mice (n=10) and controls (n=8) quantified as absolute intimal content of macrophages (Mac2 antigen immunostaining), vascular smooth muscle cells (smooth muscle α-actin, SMA immunostaining), collagen (Masson’s trichrome staining) and necrotic core (collagen-free acellular regions). Representative images of Mac2- and collagen-stained histological sections of 20% KO BMT mice are shown. b) Percentage of CD45.2+ cells within the aortic macrophage population (CD3-, Ly6g-, CD11B+, F4/80^Hi^, n = 3 pools of two aortic arches per BM genotype) and blood classical monocytes (CD3-, CD115^Hi^, Ly6g-, CD43^Lo^, Ly6c^Hi^, n=6 per BM genotype) of 20% KO BMT mice and controls, evaluated by flow cytometry. Representative CD45.1/CD45.2 plots of aortic macrophages are shown. c) Percentage of Ki-67+ proliferating cells within the aortic CD45.2+ macrophage population of 20% KO BMT mice and controls (n = 3 pools of two aortic arches per BM genotype), evaluated by flow cytometry. Representative plots are shown. d) % of S-phase cells in cultures of *Trp53*-/- and +/+ murine bone marrow-derived macrophages, evaluated by propidium iodide staining of cellular DNA content and flow cytometry; treatment with MCSF was used to induce cell cycle entry and progression in quiescent G0-synchronized macrophages (a representative experiment with macrophages from n=3 mice per genotype is shown). e, f) qPCR (e) and Western Blot (f) analyses of *Trp53* expression in cultured macrophages after MCSF mitogenic stimulation. p53 protein levels were normalized to α-tubulin. A representative blot is shown. g) Expression of cell cycle regulators *Cdkn1a*/p21^Cip1^ and *Ccnb1*/Cyclin B1 in cultured *Trp53*-/- and +/+ macrophages proliferating asynchronously (Async) or after MCSF stimulation. *p<0.05, **p<0.01, ***p<0.001.

## Discussion

This study combined exome sequencing data across two biobanks to detect somatic mutations in over 50,000 individuals and observed that the presence of CHIP was significantly associated with an increased risk of developing PAD and atherosclerosis across multiple arterial beds. Increased risk was differentially observed across CHIP driver genes with evidence of a graded relationship with CHIP VAF, with large CHIP clones conferring greater risk of disease. Lastly, analysis of p53 CHIP using a BMT murine model showed increased aortic atherosclerotic plaque among p53 CHIP carriers accompanied by expansion of plaque macrophages, supporting a direct contribution of p53-mutant hematopoietic cells to accelerated atherogenesis.

These findings permit several conclusions. First, in humans, CHIP appears to promote atherosclerosis across the entire arterial system. Previous work linked CHIP with increased risk of coronary artery disease and early-onset MI^2, 3^. We build on these findings by demonstrating that CHIP is also associated with PAD, and a composite pan-arterial atherosclerosis outcome reflective of an increased burden of atherosclerosis throughout the human arterial system. In addition, we observed suggestive CHIP associations with aneurysmal degeneration of the aorta. The genetic and epidemiologic risk factors underlying atherosclerotic occlusive disease and AAA overlap considerably^27^, and substantial evidence implicates inflammation and macrophage infiltration in driving this aortic disease^28^. The observed link between CHIP and aneurysmal disease warrants further investigation in future studies.

Second, DDR gene CHIP appears to confer an increased risk of atherosclerotic cardiovascular disease. In previous work, Jaiswal et al demonstrated an increased risk of coronary artery disease among individuals with CHIP through *DNMT3A, TET2, ASXL1,* and *JAK2* somatic driver mutations^3^. DDR CHIP is often observed among individuals following cytotoxic chemotherapy for the treatment of malignancy and has been linked to the development of therapy-related myeloid neoplasms, but limited evidence is available to inform clinical decision making regarding whether DDR CHIP carriers have increased risk of cardiovascular disease. The current study demonstrates that CHIP related to DDR-genes (*TP53, PPM1D*) confers an increased risk of developing atherosclerosis. These findings lend human genetic support to the concept that post-cytotoxic chemotherapy patients may benefit from surveillance for atherosclerotic conditions in addition to therapy-related myeloid neoplasms.

Third, CHIP related to *TP53* mutations appears to drive atherosclerosis risk via expansion of p53-deficient macrophages in occlusive plaque lesions. Previous experimental studies assessed the role of p53 in atherogenesis using mice engineered to exhibit gain or loss of function of p53 in the whole body or in specific cell types, with a variety of results^29–35^. Here we demonstrate that carrying a fraction of p53-deficient blood cells is sufficient to accelerate atherosclerosis development, as these cells have a selective advantage to expand, both in HSPCs within the BM and in macrophages within the arterial wall, leading to increased macrophage burden in the atherosclerotic plaque. Mechanistically, p53-deficient macrophage expansion seems related to increased proliferation, a major driver of macrophage burden in atherosclerotic plaques^36^. Although p53 expression is typically induced by DNA damage or other kinds of cellular stress, we found that it is expressed in resting conditions in cultured macrophages and further induced by mitogenic stimulation, suggesting a physiological role in the regulation of macrophage cell cycle progression. Accordingly, p53-deficient macrophages exhibited accelerated cell cycle kinetics. Overall, these experimental findings provide support to the notion that *TP53* CHIP mutations contribute directly to accelerated atherosclerosis and highlight major differences in the mechanisms underlying accelerated atherosclerosis in CHIP driven by mutations in *TP53* or epigenetic regulators. Previous murine studies on *Tet2* CHIP showed that TET2-deficient cells expand in bone marrow and blood, but not in the atherosclerotic plaque. In contrast, we found that p53 exhibits a double competitive advantage, expanding in both the BM and arterial wall, which suggests that small *TP53*-mutant blood clones may be sufficient to accelerate atherosclerosis development. Evaluating this possibility will require additional studies using high sensitivity sequencing strategies. Furthermore, in contrast to previous reports related to *DNMT3A/TET2* CHIP^12, 17, 18, 24, 25^, we did not observe a significant effect of p53 deficiency on the expression of the pro-inflammatory cytokines IL-1β and IL-6. These results are consistent with previous human data showing that circulating levels of pro-inflammatory cytokines are significantly increased in carriers of CHIP driven by mutations in these epigenetic regulators, but not in carriers of mutations in *TP53* or *PPM1D*^10^. These mechanistic differences between *TP53*- CHIP and *DNMT3A/TET2*-CHIP require consideration when designing preventive care strategies targeting the effects of CHIP on atherosclerosis. Thus, while the pathogenic effects of TET2-CHIP may be prevented by targeting IL-1β-driven inflammation^17^, accelerated atherosclerosis associated with *TP53* mutations may be better tackled by other strategies. Additional experimental and clinical studies should evaluate these opportunities for personalized medicine in the context of CHIP.

Lastly, a recent publication has generated a novel hypothesis regarding the relationship between CHIP and atherosclerosis^37^. In their analysis, Heyde et al. suggest that CHIP may be a symptom of the atherosclerosis trait complex (the interplay of chronic inflammation, hyperlipidemia, and arterial plaque), rather than a causal risk factor, based on murine and human observations that atherosclerosis may accelerate HSC proliferation and somatic evolution. While their study engenders an intriguing hypothesis, there are several observations in the literature and in our current study that we believe do not support this interpretation. Firstly, in the Heyde et al. study, *Ldlr*-null mice irradiated and transplanted with bone marrow of *Tet2* -/- mice that were fed an atherogenic diet demonstrated greater *Tet2* -/- neutrophil and monocyte expansion than similar mice fed normal chow. However, in marked contrast, we previously showed that *Tet2*-/- cells expand similarly when *Ldlr*-null mice are fed either a high cholesterol diet or normal chow^17^. Furthermore, in our prior work, the expansion of *Tet2*-/- cells was completely unaffected by treatment with a pharmacological NLRP3 inhibitor, even though this inhibitor led to a 2-fold decrease in atherosclerotic plaque size^17^. Secondly, in our current analysis of the pan-arterial atherosclerosis endpoint, CHIP was associated with incident atherosclerotic events among a pool of individuals without any history of atherosclerosis at baseline - suggesting that CHIP precedes clinically significant occlusive disease. Thirdly, Heyde et al. state that the uniform hazard ratios observed for the CHIP-coronary artery disease association provide supportive evidence for the notion that atherosclerotic disease precedes the development of clonal hematopoiesis. However, in our study we identify significant heterogeneity of incident PAD effect sizes when stratifying by CHIP genes, as well as a dose-response relationship between CHIP clone size (or VAF) and incidence of atherosclerosis, suggesting that CHIP genes vary in their effect on incident PAD risk, and that expanded CHIP variants have a stronger influence on future atherosclerotic risk. Thus, given the 1) association with incident atherosclerotic disease, 2) dose-response relationship with CHIP clone size and heterogeneity by CHIP gene, and 3) that *TP53*, *TET2*, and *JAK2*^38^ appear to drive atherosclerosis through disparate mechanisms, clonal hematopoiesis acting as a causal risk factor appears to be a more parsimonious explanation to unify these findings.

Several limitations merit mention. First, our PAD and cardiovascular disease phenotypes are based on EHR data and may result in misclassification of case status. Such misclassification would likely reduce statistical power for discovery and on average bias results toward the null. Second, selection bias from differential loss-of-follow up, volunteer bias, and missingness in covariates may be present given the nature of the genetic biobanks used in this study. Third, while we maximized the number of participants in our analysis of CHIP and additional atherosclerotic diseases, it may still have been underpowered for certain phenotypes. Lastly, the cohorts in these studies are largely of European ancestry; while prior reports have not observed significant phenotypic differences in CHIP associations by ancestry^5, 12^, further epidemiological studies in ethnically diverse individuals are required to ensure that this is the case for PAD as well.

In conclusion, CHIP is associated with incident atherosclerosis in multiple vascular beds, with murine evidence of increased plaque among *TP53* CHIP carriers through an expansion of plaque macrophages (**Figure 6**). The observations presented here expand insight into CHIP-mediated atherosclerosis. The novel finding that DDR gene CHIP plays a particular role in PAD raises new questions regarding the mechanisms of regional heterogeneity of atherosclerotic involvement in face of systemic exposure to traditional risk factors such as dyslipidemia, hypertension, and smoking.

**Figure 6.**
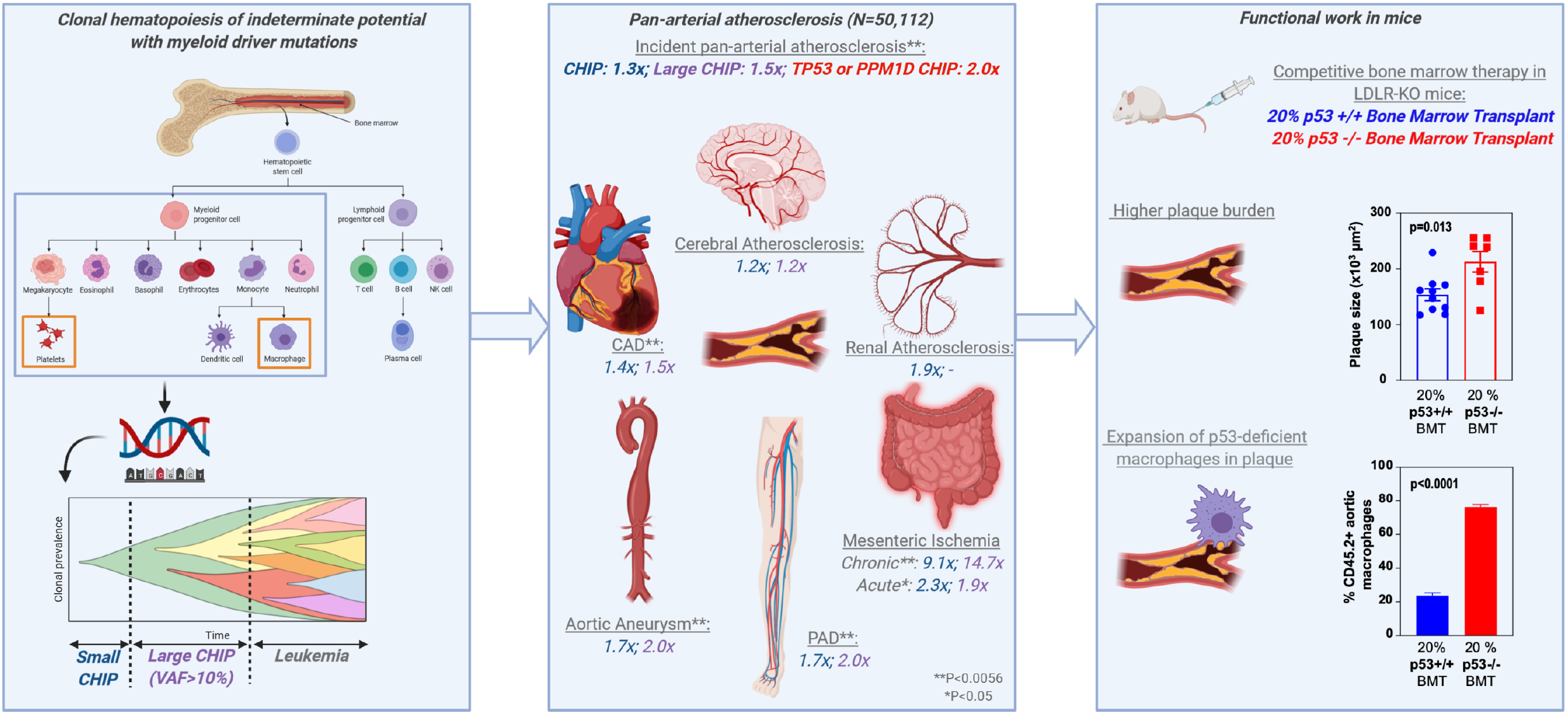
In this study, we assessed the association of clonal hematopoiesis of indeterminate potential (CHIP) with myeloid driver mutations with pan-arterial atherosclerosis. CHIP is a category of age-related somatic variants which are associated with incident leukemia and thought to be implicated in atherosclerosis primarily by altering macrophage function and promoting thrombosis. CHIP clones can be characterized by the fraction of blood cells carrying the clone, referred to as the variant allele fraction (VAF); here we categorized large CHIP clones as variants with VAF>10%. Across 50,112 individuals from the UK Biobank and Mass-General Brigham Biobank, we observed that CHIP is associated with increased risk of incident peripheral and pan-arterial atherosclerosis, with stronger effects conferred by large CHIP clones (HR 1.5x). In addition, we observed and a novel associations for *TP53* and *PPM1D* CHIP (HR 2.0x). CHIP was found to be individually associated with a variety of atherosclerotic conditions, with Bonferroni-significant associations (double-starred, **) identified for peripheral artery disease (PAD), coronary artery disease (CAD), aortic aneurysm, and chronic mesenteric ischemia. HR for CHIP are displayed in blue and for large CHIP in purple. Functional analysis was performed to further investigate the observed TP53-PAD association. *Ldlr*-KO 20% p53 -/- bone-marrow transplanted mice had a significant increase in plaque size, with significant expansion of p53-deficient macrophages in plaque (P<0.001) at 12 weeks.

## Data Availability

UKB individual-level data are available for request by application (https://www.ukbiobank.ac.uk). Individual-level MGBB data are available from https://personalizedmedicine.partners.org/Biobank/Default.aspx, but restrictions apply to the availability of these data, which were used under IRB approval for the current study, and so are not publicly available. The present article includes all other data generated or analyzed during this study.

## Acknowledgments

Thanks to the participants and staff of the UK Biobank and Mass General Brigham Biobank. UK Biobank analyses were conducted using Application 7089. Thanks to Dr. Zeyan Liew and teaching fellow Jiajun Lu, the instructors of the causal inference course at Yale.

## Abbreviations

CHIP: clonal hematopoiesis of indeterminate potential
PAD: peripheral artery disease

---

**Supplementary Figure I.**
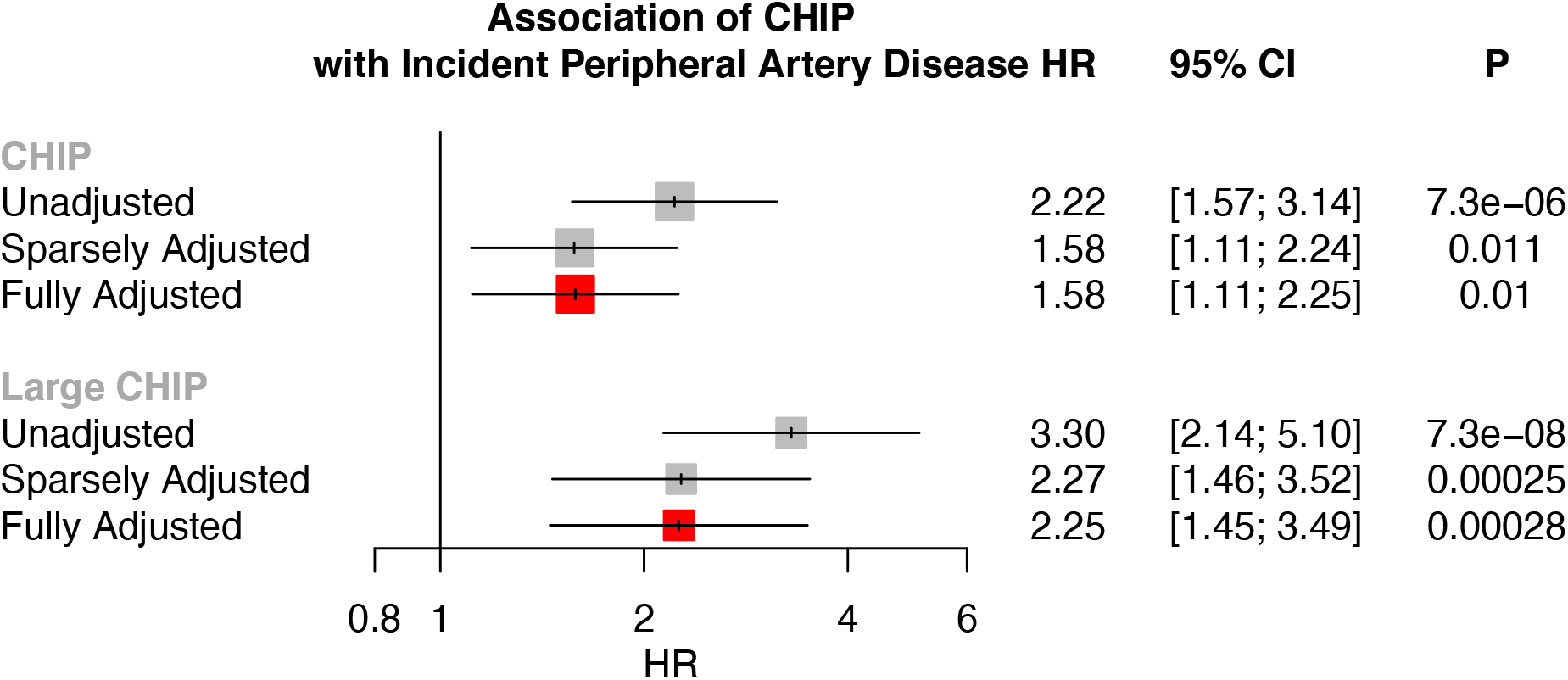
Association of CHIP and Large CHIP (variant allele fraction>10%) with PAD in the UKB under 1) unadjusted, 2) sparsely adjusted, and 3) fully adjusted models, where sparsely adjusted refers to the following covariates: age, age^2^, sex, smoking status, Townsend deprivation index, and the first ten principal components of genetic ancestry, and the fully adjusted model additionally includes normalized BMI, prevalent hypertension, hyperlipidemia, and type 2 diabetes as covariates. Given the minimal difference between the sparsely adjusted and fully adjusted model, the sparsely adjusted model was moved forward for use in analysis.

**Supplementary Figure II.**
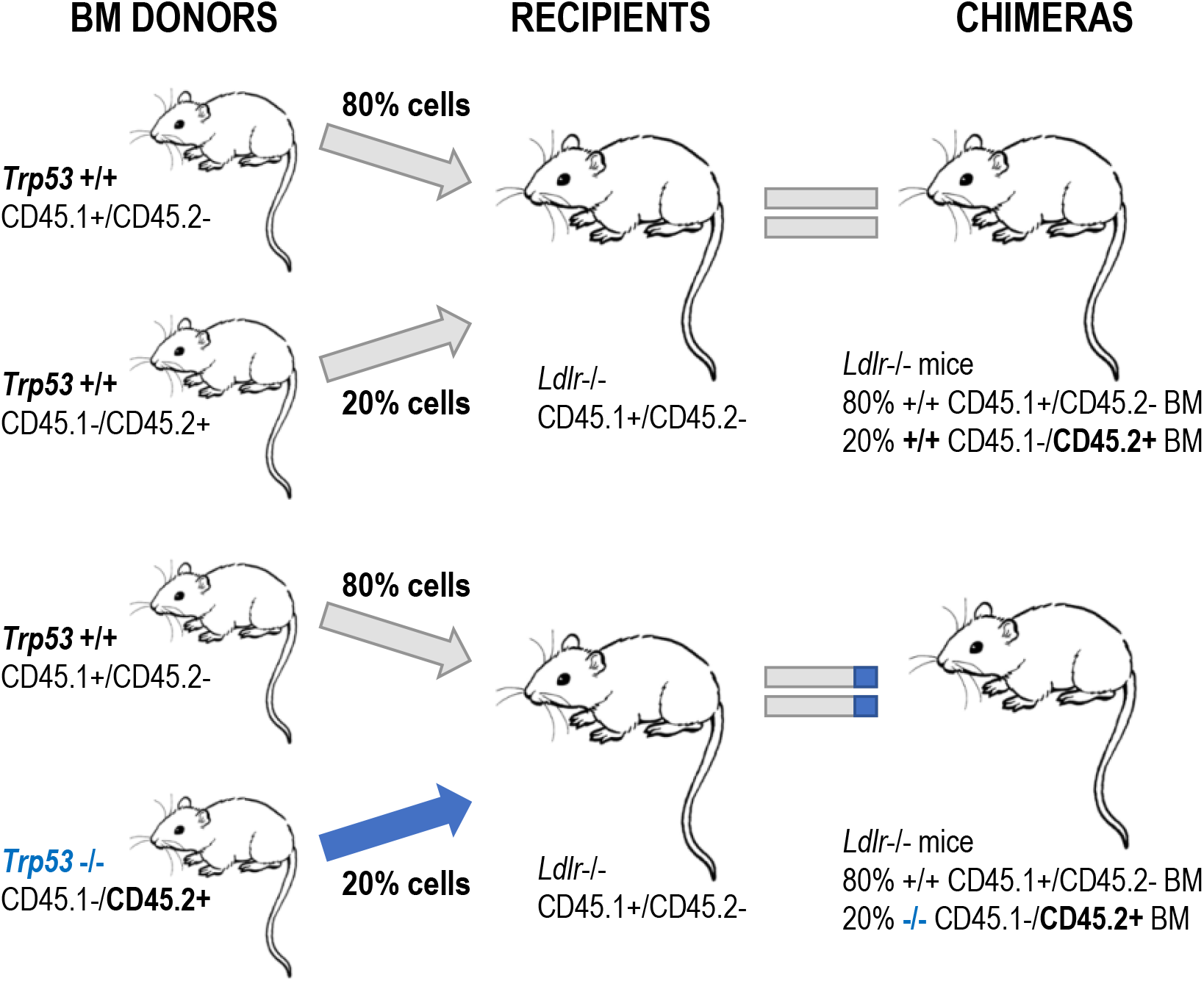
Competitive bone marrow transplantation approach to generate a murine model of p53 CHIP

**Supplementary Figure III.**
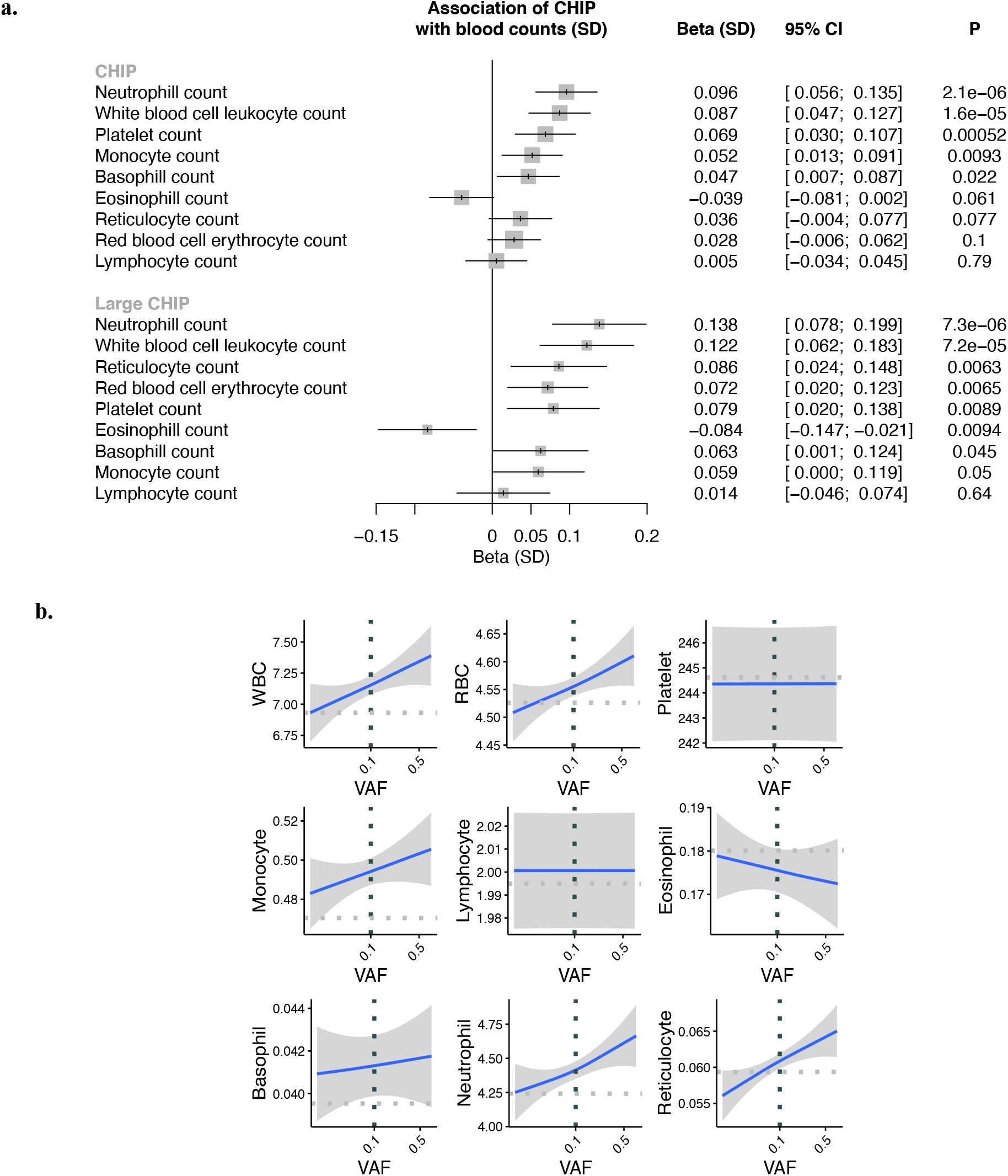
Association of CHIP with blood counts among individuals without prevalent hematologic malignancy in the UK Biobank. Blood counts were acquired at time of blood draw for whole exome sequencing. a) Association of CHIP and Large CHIP with normalized blood counts (SD). Associations are adjusted for age, age^2^, sex, smoking status, and the first ten principal components of genetic ancestry. b) Association of CHIP variant allele frequency (VAF) with blood counts (in units of 10^9 cells/L). The gray horizontal dotted lines reflect average counts across non-CHIP carriers. The vertical black dotted line reflects the cutoff VAF for Large CHIP (VAF>0.1). CHIP = clonal hematopoiesis of indeterminate potential; VAF = variant allele fraction

**Supplementary Figure IV:**
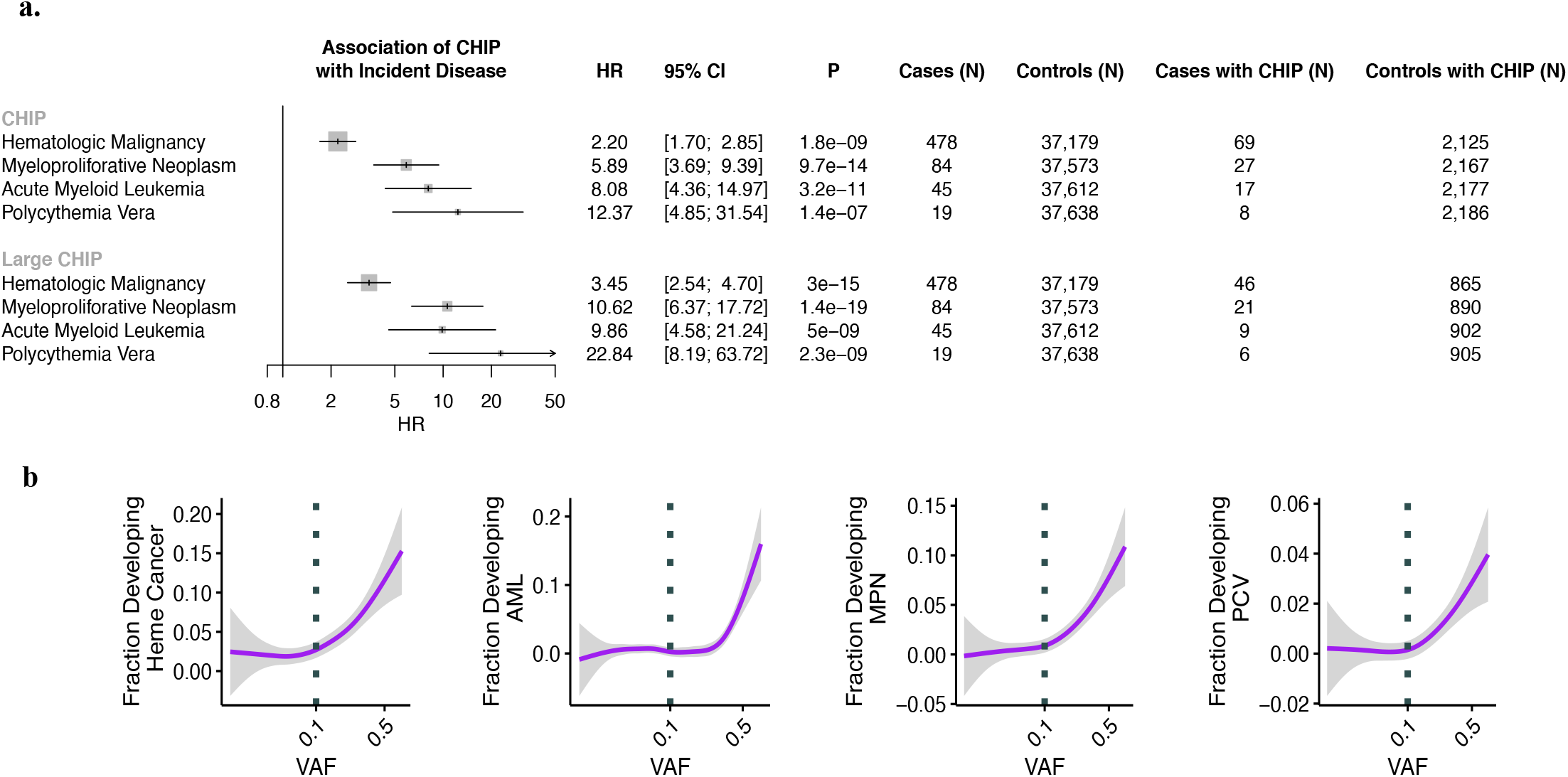
Association of CHIP (a) and VAF (b) with incident hematologic malignancy among individuals without prevalent hematological malignancy in the UK Biobank. Associations are adjusted for age, age^2^, sex, smoking status, Townsend deprivation index, and the first ten principal components of genetic ancestry. CHIP = clonal hematopoiesis of indeterminate potential; VAF = variant allele fraction

**Supplementary Figure V:**
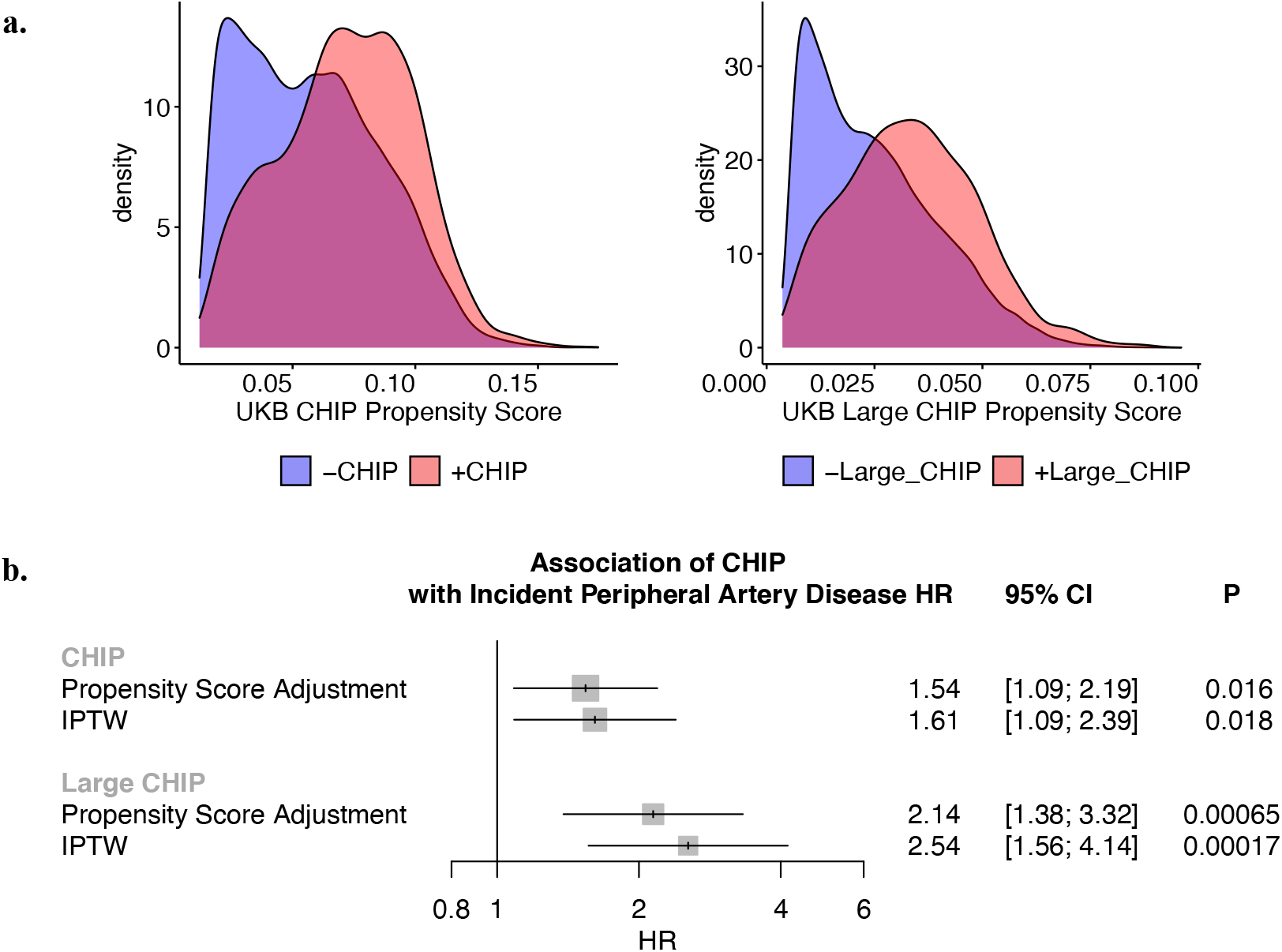
Epidemiological causal inference analysis for CHIP on incident peripheral artery disease in the UK Biobank. **a)** Propensity scores by CHIP and Large CHIP status in the UKB. **b)** Propensity score adjustment and stabilized inverse probability treatment weighting (IPTW) for the CHIP and Large CHIP association with incident PAD in the UKB. CHIP = clonal hematopoiesis of indeterminate potential; VAF = variant allele fraction; PAD = peripheral artery disease

**Supplementary Figure VI:**
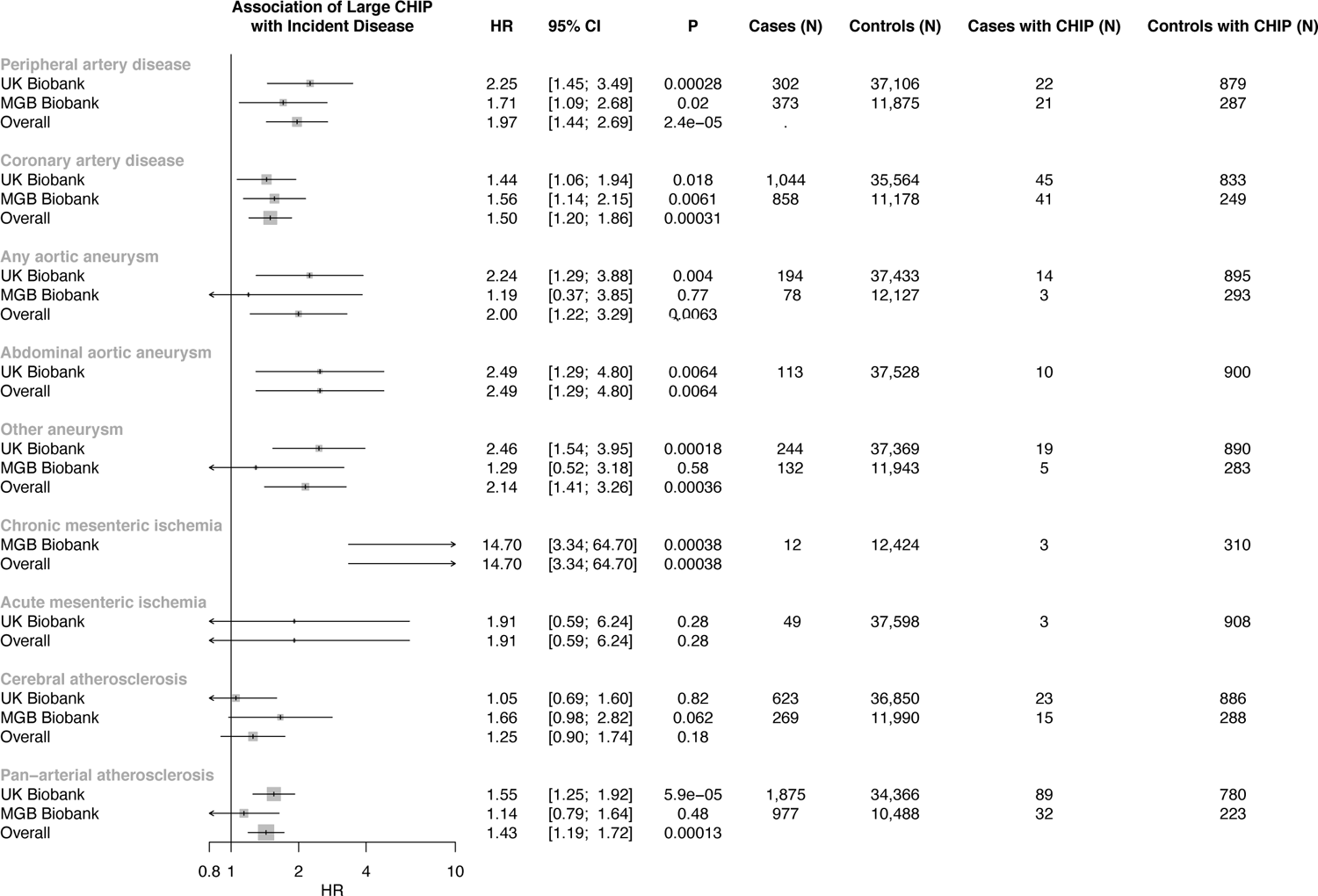
Association of Large CHIP (VAF>10%) with incident pan-arterial atherosclerosis, combined across peripheral artery disease, coronary artery disease, aneurysms, chronic and acute mesenteric ischemia, cerebral atherosclerosis, and renal artery stenosis. CHIP = clonal hematopoiesis of indeterminate potential; VAF = variant allele fraction

**Supplementary Figure VII:**
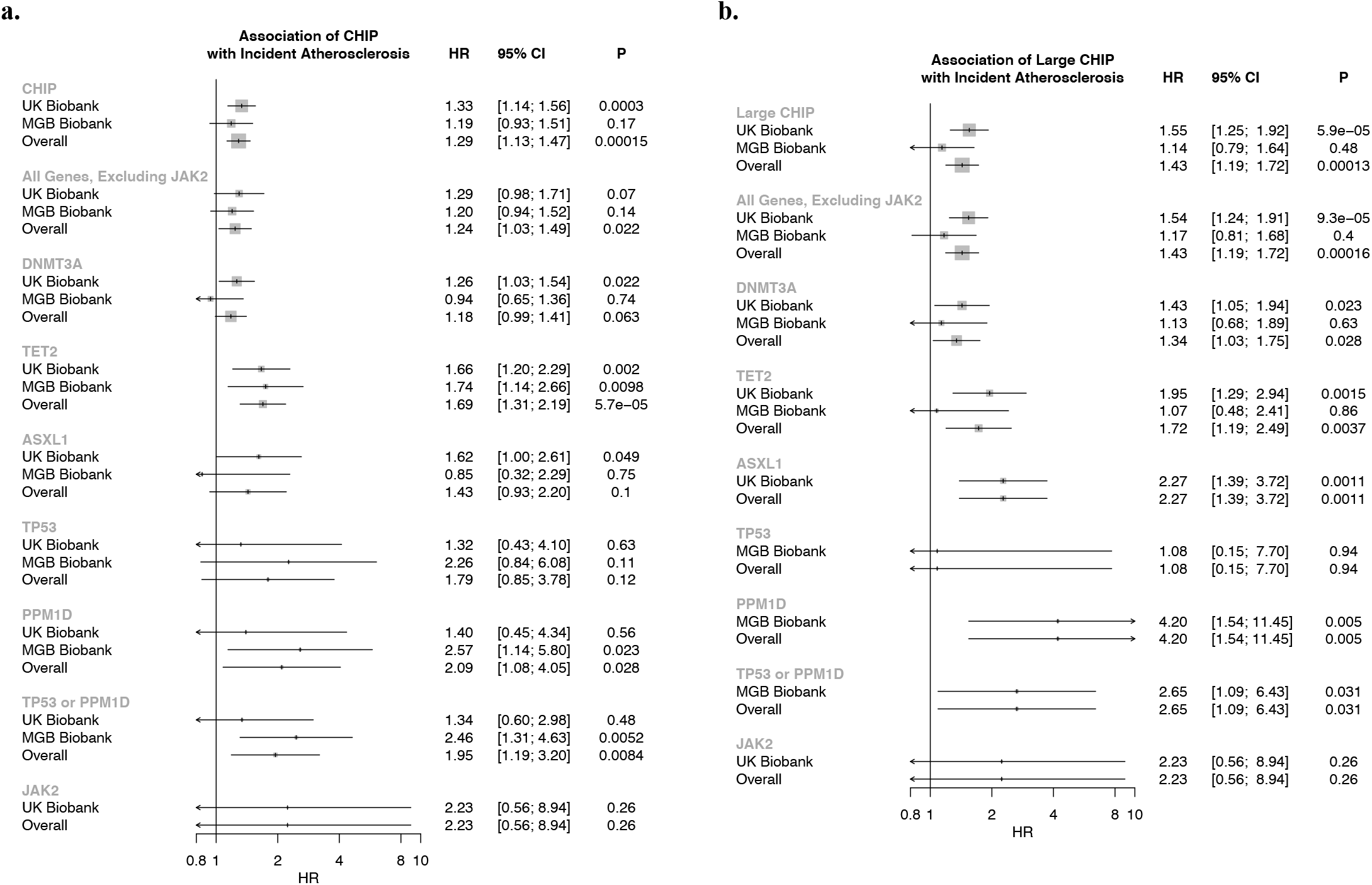
Association of **a)** CHIP and **b)** Large CHIP genes with incident pan-arterial atherosclerosis, combined across peripheral artery disease, coronary artery disease, aneurysms, chronic and acute mesenteric ischemia, cerebral atherosclerosis, and renal artery stenosis. CHIP = clonal hematopoiesis of indeterminate potential; VAF = variant allele fraction

**Supplementary Figure VIII:**
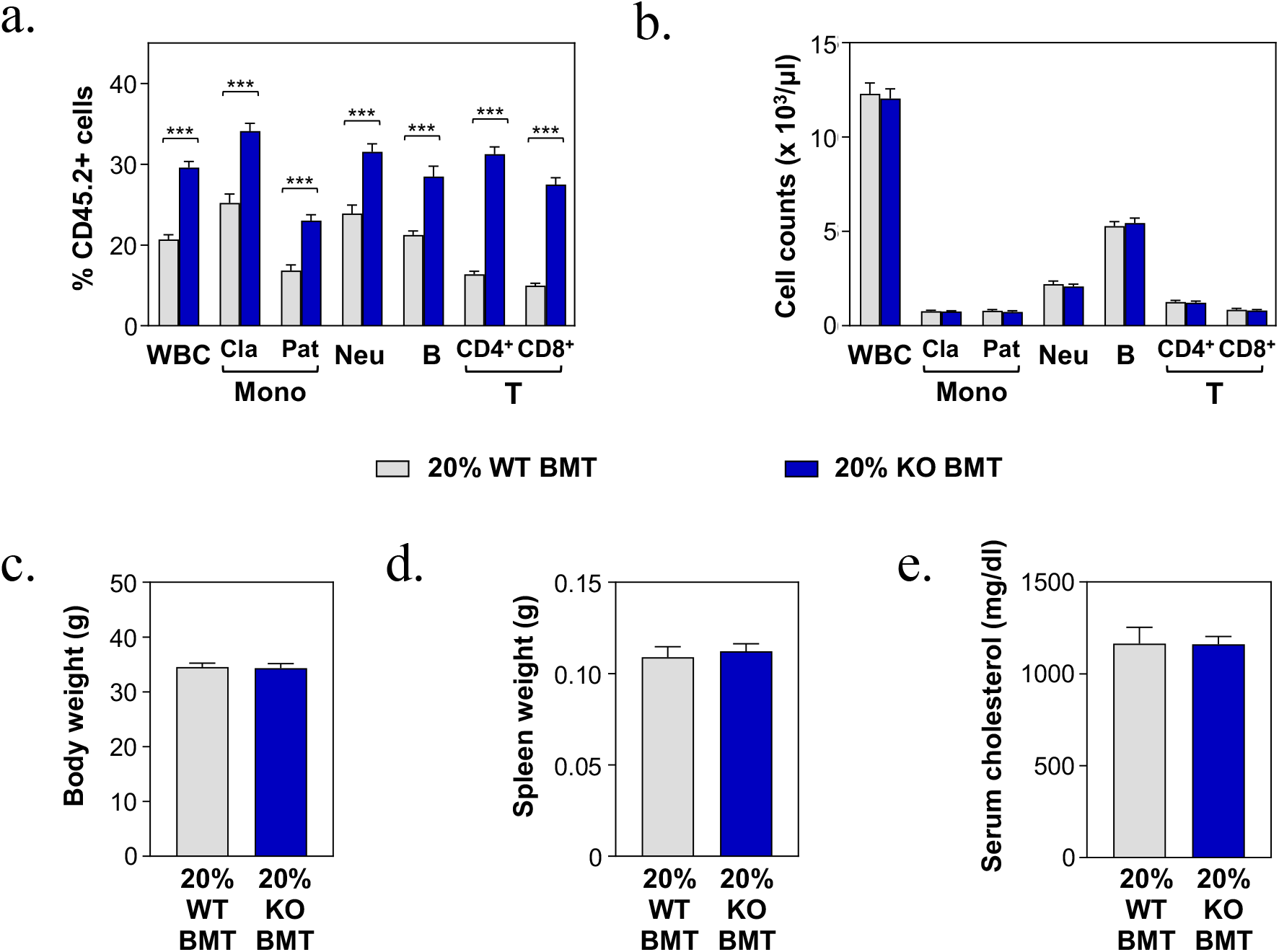
20% KO-BMT male mice and 20% WT-BMT controls were fed a high-fat/high-cholesterol (HF/HC) diet for 9 weeks, starting 4 weeks after BMT. a) Percentage of CD45.2+ cells in different white blood cell (WBC) lineages in peripheral blood, evaluated by flow cytometry (***p<0.001). b) Absolute counts of main WBC sub-populations in peripheral blood, evaluated by flow cytometry. c) Body weight. d) Spleen weight. e) Total cholesterol level in serum, evaluated by enzymatic methods.

**Supplementary Figure IX.**
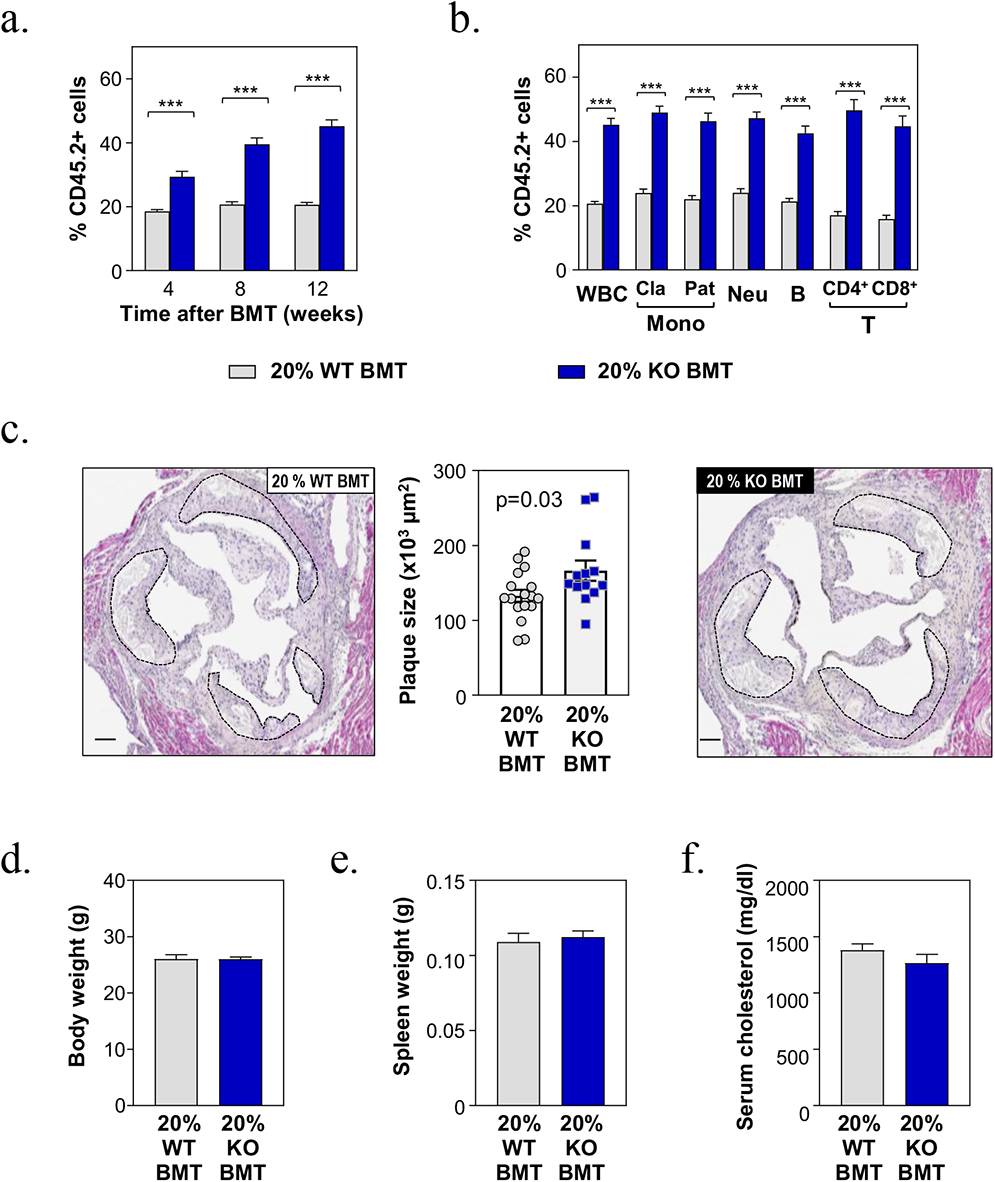
20% KO-BMT female mice (n=13) and 20% WT-BMT controls (n=17) were fed a high-fat/high-cholesterol (HF/HC) diet for 9 weeks, starting 4 weeks after BMT. a) Percentage of CD45.2+ in white blood cells at different timepoints, evaluated by flow cytometry (***p<0.001). b) Percentage of CD45.2+ cells in different white blood cell (WBC) lineages in peripheral blood after 9 weeks on HF/HC diet (13 weeks post-BMT), evaluated by flow cytometry (***p<0.001). c) Aortic root plaque size. Representative images of hematoxylin and eosin-stained sections are shown; atherosclerotic plaques are delineated by dashed lines. Scale bars, 100 µm. d) Body weight. e) Spleen weight. f) Total cholesterol level in serum, evaluated by enzymatic methods.

**Supplementary Figure X.**
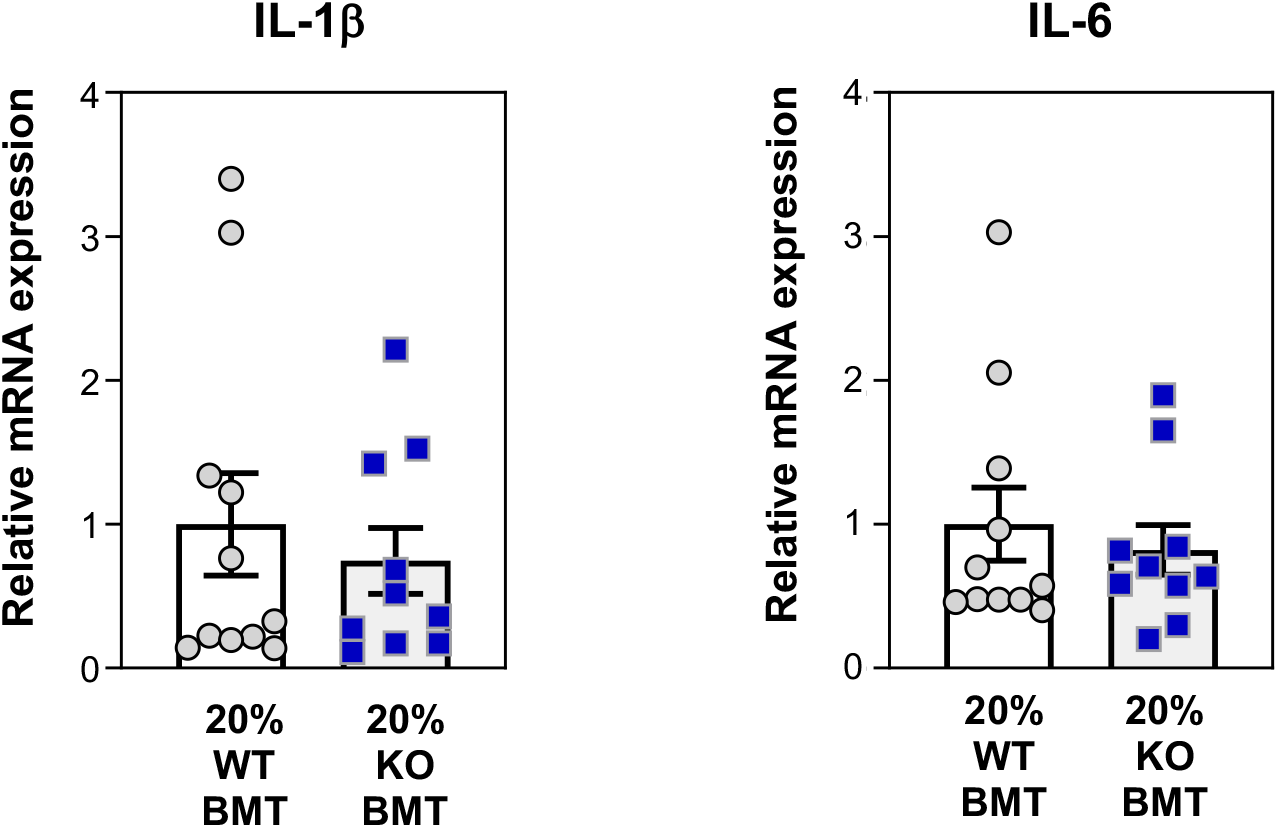
Aortic arch samples were obtained from HF/HC-fed 20% WT-BMT mice (n=11) or 20% KO-BMT mice (n=10) and gene expression was analyzed by qPCR analysis.

