## Supplementary Tables for "*TP53-*mediated clonal hematopoiesis confers increased risk for incident peripheral artery disease"

**Supplementary Methods**

**Epidemiologic causal inference methods used to assess the CHIP-PAD association:**

The propensity score is the probability of treatment assignment (i.e. clonal hematopoiesis of indeterminate potential [CHIP] or large CHIP) conditional on the minimally sufficient set of confounders. Using a propensity score as a covariate allows observational studies to mimic some of the characteristics of a randomized controlled trial in the sense that conditional on the propensity score, the distribution of covariates will be similar between the CHIP and the non-CHIP carriers. In our scenario, the propensity score will help assess the ‘dimensionality problem’ in that it will reduce the number of covariates from 15 covariates in the minimally sufficient set to 1. Here, the propensity score is calculated using multivariable logistic regression where the outcome was CHIP and covariates consisted of the 15 covariates in the minimally sufficient set. The propensity score was then predicted for all samples in the UK Biobank for CHIP and, separately, for large CHIP.

Stabilized Inverse probability of treatment weighting (IPTW) uses weights based on the propensity score to create a pseudo-population in which the distribution of measured covariates is independent of CHIP status, i.e. removing the arrow from the confounders to the exposure. The conditional exchangeability assumption made in IPTW is that if the set of covariates are sufficient to block all backdoor paths from CHIP to peripheral artery disease (PAD), then all confounding is eliminated in the pseudo-population and thus the association between CHIP and PAD in the synthetic population is an estimate of the causal effect. To estimate the IPTW stabilized weight ($SW$) for each individual in the UKB, we utilized the following equation for CHIP carriers and controls, where $PS$ reflects the propensity score for that individual:

$${SW}^{CHIP+}=\frac{\Pr\left( CHIP=1 \right)}{\Pr\left( CHIP=1 \right|covariates)}=\frac{\Pr\left( CHIP=1 \right)}{PS}$$

$${SW}^{CHIP-}=\frac{1-Pr \left( CHIP=1 \right)}{\Pr\left( CHIP=0 \right|covariates)}=\frac{\Pr\left( CHIP=0 \right)}{PS}$$

Supplementary Tables

Tables I-II are provided in the separate excel sheet

**Supplementary Table III -** CHIP gene carrier count by cohort. Splicing Factor Mutations refer to the following CHIP genes: *LUC7L2, PRPF8, SF3B1, SRSF2, U2AF1,* and *ZRSR2.* Large CHIP refers to mutations with variant allele frequency > 10%.

|  | UKBB (N=37,657) | | MGBB (N=12,465) | |
| --- | --- | --- | --- | --- |
|  | All CHIP | Large CHIP | All CHIP | Large CHIP |
| CHIP (%) | 2194 (5.8) | 911 (2.4) | 657 (5.3) | 314 (2.5) |
| >1 CHIP Mutation (%) | 191 (0.5) | 70 (0.2) | 55 (0.4) | 16 (0.1) |
| DNMT3A (%) | 1401 (3.8) | 489 (1.4) | 311 (2.6) | 144 (1.2) |
| TET2 (%) | 347 (1.0) | 181 (0.5) | 132 (1.1) | 61 (0.5) |
| JAK2 (%) | 17 (0.0) | 17 (0.0) | 5 (0.0) | 5 (0.0) |
| ASXL1 (%) | 152 (0.4) | 100 (0.3) | 47 (0.4) | 21 (0.2) |
| Splicing Factor Mutation (%) | 49 (0.1) | 28 (0.1) | 17 (0.1) | 8 (0.1) |
| TP53 (%) | 36 (0.1) | 11 (0.0) | 20 (0.2) | 12 (0.1) |
| PPM1D (%) | 32 (0.1) | 12 (0.0) | 32 (0.3) | 13 (0.1) |
| TP53 or PPM1D (%) | 68 (0.2) | 23 (0.1) | 52 (0.4) | 25 (0.2) |

**Supplementary Table IV -** Demographic and clinical characteristics for CHIP carriers and controls in the UK and Mass General Brigham Biobanks. P-values reflect chi-square tests comparing CHIP carriers to controls across each phenotypic category.

|  | UK Biobank | | | | | MGB Biobank | | | | | |
| --- | --- | --- | --- | --- | --- | --- | --- | --- | --- | --- | --- |
|  | **-CHIP** | **+CHIP** | | **p** | | **-CHIP** | **+CHIP** | | | **p** | |
| n | 35463 | 2194 |  | | | 11808 | 657 | | | |  |
| age (mean (SD)) | 56.81 (7.84) | 60.59 (6.57) | <0.001 | | | 46.13 (14.65) | 60.12 (12.05) | | | | <0.001 |
| Sex = Male (%) | 16379 (46.2) | 1042 (47.5) | 0.242 | | | 4937 (41.8) | 304 (46.3) | | | | 0.027 |
| Race (%) |  |  | NA | | |  |  | | | | 0.002 |
| White | 35463 (100.0) | 2194 (100.0) | | | | 9449 (80.0) | 566 (86.1) |  | | | |
| Black |  |  | | |  | 723 (6.1) | 27 (4.1) | |  | | |
| Asian |  |  | | |  | 465 (3.9) | 19 (2.9) | |  | | |
| Other |  |  | | |  | 474 (4.0) | 12 (1.8) | |  | | |
| Unknown |  |  | | |  | 697 (5.9) | 33 (5.0) | |  | | |
| Smoking Status (%) |  |  | | | <0.001 |  |  | | <0.001 | | |
| Current | 3027 (8.5) | 220 (10.0) | | |  | 288 (2.4) | 17 (2.6) | |  | | |
| Previous | 12664 (35.7) | 900 (41.0) | | |  | 3662 (31.0) | 261 (39.7) | |  | | |
| Never | 19772 (55.8) | 1074 (49.0) | | |  | 7183 (60.8) | 351 (53.4) | |  | | |
| Alcohol intake (drinks in last 4wk) (mean (SD)) | 11.37 (9.89) | 11.67 (10.11) | | | 0.156 |  |  | |  | | |
| Exercise frequency (days in last 4wk) (mean (SD)) | 8.34 (6.38) | 8.45 (6.43) | | | 0.591 |  |  | |  | | |
| Townsend Deprivation Index (mean (SD)) | -1.55 (2.81) | -1.65 (2.75) | | | 0.137 |  |  | |  | | |
| Significant life stressor in last 2y (%) | 16915 (47.8) | 1030 (47.1) | | | 0.535 |  |  | |  | | |
| Handfulls of sweets/day (mean (SD)) | 1.09 (1.17) | 0.93 (1.19) | | | 0.399 |  |  | |  | | |
| Vegetable servings/day (mean (SD)) | 1.08 (0.54) | 1.02 (0.49) | | | 0.283 |  |  | |  | | |
| BMI (mean (SD)) | 27.39 (4.76) | 27.48 (4.55) | | | 0.414 | 28.14 (6.35) | 28.55 (6.33) | | 0.132 | | |
| Prevalent Type 2 Diabetes Mellitus (%) | 956 (2.7) | 70 (3.2) | | | 0.189 | 505 (4.3) | 40 (6.1) | | 0.035 | | |
| Prevalent Coronary Artery Disease (%) | 2040 (5.8) | 171 (7.8) | | | <0.001 | 378 (3.2) | 42 (6.4) | | <0.001 | | |
| Prevalent Hypertension (%) | 10650 (30.0) | 782 (35.6) | | | <0.001 | 1893 (16.0) | 193 (29.4) | | <0.001 | | |
| Prevalent Hypercholesterolemia (%) | 6159 (17.4) | 448 (20.4) | | | <0.001 | 1739 (14.7) | 172 (26.2) | | <0.001 | | |

**Supplementary Table V -** Breakdown of composite incident atherosclerosis events by disease in the UK Biobank and MGB Biobank.

|  | **UK Biobank** | | | **MGB Biobank** | | |
| --- | --- | --- | --- | --- | --- | --- |
|  | **No CHIP** | **CHIP** | **p** | **No CHIP** | **CHIP** | **p** |
| Composite Atherosclerosis (%) | 1697 (100.0) | 178 (100.0) | NA | 878 (100.0) | 75 (100.0) | NA |
| Coronary Artery Disease (%) | 917 (54.0) | 94 (52.8) | 0.815 | 666 (75.9) | 59 (78.7) | 0.684 |
| Peripheral Artery Disease (%) | 224 (13.2) | 33 (18.5) | 0.063 | 234 (26.7) | 19 (25.3) | 0.911 |
| Cerebral Atherosclerosis (%) | 526 (31.0) | 39 (21.9) | 0.015 | 158 (18.0) | 19 (25.3) | 0.157 |
| Abdominal Aortic Aneurysm (%) | 76 (4.5) | 13 (7.3) | 0.133 | 18 (2.1) | 0 (0.0) | 0.418 |
| Aortic Aneurysm (%) | 143 (8.4) | 24 (13.5) | 0.034 | 55 (6.3) | 4 (5.3) | 0.943 |
| Other Aneurysm (%) | 180 (10.6) | 30 (16.9) | 0.017 | 99 (11.3) | 8 (10.7) | 1 |
| Chronic Mesenteric Ischemia (%) | 3 (0.2) | 0 (0.0) | 1 | 6 (0.7) | 1 (1.3) | 1 |
| Acute Mesenteric Ischemia (%) | 37 (2.2) | 8 (4.5) | 0.097 | 1 (0.1) | 0 (0.0) | 1 |
| Renal Artery Atherosclerosis (%) | 1 (0.1) | 0 (0.0) | 1 | 14 (1.6) | 0 (0.0) | 0.547 |
